# Strategy and performance evaluation of low-frequency variant calling for SARS-CoV-2 in wastewater using targeted deep Illumina sequencing

**DOI:** 10.1101/2021.07.02.21259923

**Authors:** Laura A. E. Van Poelvoorde, Thomas Delcourt, Wim Coucke, Philippe Herman, Sigrid C. J. De Keersmaecker, Xavier Saelens, Nancy Roosens, Kevin Vanneste

**Author notes:** Equal first-author contribution (Laura A. E. Van Poelvoorde, Thomas Delcourt). Equal last-author contribution (Nancy Roosens, Kevin Vanneste). Corresponding author during submission process: Laura Van Poelvoorde, Corresponding author post-publication: Kevin Vanneste.

## Abstract

The ongoing COVID-19 pandemic, caused by SARS-CoV-2, constitutes a tremendous global health issue. Continuous monitoring of the virus has become a cornerstone to make rational decisions on implementing societal and sanitary measures to curtail the virus spread. Additionally, emerging SARS-CoV-2 variants have increased the need for genomic surveillance to detect particular strains because of their potentially increased transmissibility, pathogenicity and immune escape. Targeted SARS-CoV-2 sequencing of wastewater has been explored as an epidemiological surveillance method for the competent authorities. Few quality criteria are however available when sequencing wastewater samples, and those available typically only pertain to constructing the consensus genome sequence. Multiple variants circulating in the population can however be simultaneously present in wastewater samples. The performance, including detection and quantification of low-abundant variants, of whole genome sequencing (WGS) of SARS-CoV-2 in wastewater samples remains largely unknown. Here, we evaluated the detection and quantification of mutations present at low abundances using the SARS-CoV-2 lineage B.1.1.7 (alpha variant) defining mutations as a case study. Real sequencing data were *in silico* modified by introducing mutations of interest into raw wild-type sequencing data, or by mixing wild-type and mutant raw sequencing data, to mimic wastewater samples subjected to WGS using a tiling amplicon-based targeted metagenomics approach and Illumina sequencing. As anticipated, higher variation, lower sensitivity and more false negatives, were observed at lower coverages and allelic frequencies. We found that detection of all low-frequency variants at an abundance of 10%, 5%, 3% and 1%, requires at least a sequencing coverage of 250X, 500X, 1500X and 10,000X, respectively. Although increasing variability of estimated allelic frequencies at decreasing coverages and lower allelic frequencies was observed, its impact on reliable quantification was limited. This study provides a highly sensitive low-frequency variant detection approach, which is publicly available at https://galaxy.sciensano.be, and specific recommendations for minimum sequencing coverages to detect clade-defining mutations at specific allelic frequencies.

## 1 Introduction

Severe acute respiratory syndrome coronavirus 2 (SARS-CoV-2) is the causative agent of the ongoing COVID-19 pandemic [1]. To limit the spread of disease, governments were forced to take drastic measures due to the high potential for human-to-human transmission and the lack of immunity in the population [2]. SARS-CoV-2 spreads very easily during close person-to-person contact [3]. Consequently, the individual diagnostic testing for SARS-CoV-2 on respiratory samples using reverse transcription quantitative polymerase chain reaction (RT-qPCR) is essential for the diagnosis of patients presenting COVID-19 symptoms for appropriate clinical treatment and isolation, as well as for tracing potential contact transmissions, including asymptomatic individuals. Systematic individual SARS-CoV-2 diagnostics are also used to test certain population cohorts, such as primary caregivers, to avoid transmission of the virus to vulnerable people, such as the elderly.

Data from individual diagnostics are also collected and analysed for surveillance by National Reference Centres to assist governments to monitor the epidemiological situation. The efficiency of this strategy for epidemiological monitoring depends greatly on the extent of testing the complete population. Additionally, it may be biased by the willingness of individuals, covering all population ages, to get tested, whether individuals are aware of being infected, and visitors to a certain country not always being included in the testing strategy. Moreover, despite having a relatively low per-sample cost, the high volume of required tests incurs substantial costs for public health systems for which testing capacities can be exceeded during periods of intense circulation of the virus [4]. The detection of newly emerging SARS-CoV-2 strains may be delayed by the lack of testing during such periods. As SARS-CoV-2 viral particles and mRNA have been isolated from faeces of COVID-19 patients [5, 6], monitoring of wastewater for SARS-CoV-2 has been explored as a complementary and independent alternative for epidemiological surveillance for the competent authorities [7]. Various studies have observed an association between an increase in reported COVID-19 cases and an increase of SARS-CoV-2 RNA concentrations in wastewater [8–10]. Wastewater-based monitoring could therefore be a cost-effective, non-invasive, easy to collect, and unbiased approach to track circulating virus strains in a community [11]. Compared to clinical surveillance, wastewater surveillance could also provide opportunities to estimate the prevalence of the virus and assess its geographic distribution and genetic diversity [12, 13], and can be used as a non-invasive early-warning system for alerting public health authorities to the potential (re-)emergence of COVID-19 infections [14]. Alternatively, the absence of the virus in wastewater surveillance could indicate that an area can be considered at low risk for SARS-CoV-2 infections [7].

Although the mutation rate of SARS-CoV-2 is estimated as being low compared to other RNA viruses [15], several new variants carrying multiple mutations have already emerged. Some of these variants are characterized by a potential enhanced transmissibility, and can cause more severe infections and/or potential vaccine escape [16–20]. Consequently, monitoring current and potential future variants, is crucial to control the epidemic by taking timely measures because these variants can affect epidemiological dynamics, vaccine effectiveness and disease burden.

To monitor SARS-CoV-2 variants, RT-qPCR methods were designed to detect a selection of the mutations that define specific variants of concern (VOCs). VOCs are however defined by a combination of multiple mutations and only few mutations can be targeted by RT-qPCR assays, but many VOCs are characterized by a high number of specific mutations. This approach is also not sustainable because it is likely that the ongoing vaccination and increased herd immunity will result in the selection of new mutations and emergence of new VOCs [21], as has been observed with other viruses [22, 23]. Since only a few mutations can be targeted by a RT-qPCR assay, an additional step of whole genome sequencing (WGS) is required to fully confirm the variant’s sequence [24].

WGS has been used to understand the viral evolution, epidemiology and impact of SARS-CoV-2 resulting in, as of July 2021, more than 2,000,000 publically available SARS-CoV-2 genome sequences, mainly derived from respiratory samples that are frequently submitted to the Global Initiative on Sharing Avian Influenza Data (GISAID) database [25]. Most of these sequences were obtained using amplicon sequencing in combination with the Illumina or Nanopore technology, with Illumina still being the most commonly used method [25, 26]. This large amount of genomes allows reliable detection of variants based on the consensus genome sequence in patient samples [27–30]. The European Centre for Disease Prevention and Control (ECDC) has defined several quality criteria for clinical samples depending on the application. For most genomic surveillance objectives, a consensus sequence of the (near-)complete genome is sufficient and a minimal read length of 100 bp and minimal coverage of 10X across more than 95% of the genome is recommended. To reliably trace direct transmission and/or reinfection, a higher sequencing coverage of 500X across more than 95% of the genome is recommended for determining low-frequency variants (LFV) that can significantly contribute to the evidence for reinfection or direct transmission. In-depth genome analysis, including recombination, rearrangement, haplotype reconstruction and large insertions and deletions (indel) detection, should be investigated using long-read sequencing technologies with a recommended read length of minimally 1000 bp and a sequencing coverage of 500X across more than 95% of the genome [31]. Due to the high cost of sequencing large quantities of samples from individual patient, samples that tested positive for a selection of mutations related to VOCs using RT-qPCR and have a sufficiently high viral load are typically sequenced. Consequently, only a subset of all circulating variants is detected during routine clinical surveillance. Since wastewater samples contain both SARS-CoV-2 RNA from symptomatic and asymptomatic individuals, sequencing wastewater samples can provide a more comprehensive picture of the genomic diversity of SARS-CoV-2 circulating in the population compared to individual clinical testing and sequencing. Wastewater surveillance of SARS-CoV-2 may therefore be of considerable added value for SARS-CoV-2 genomic surveillance by providing a cost-effective, rapid and reliable source of information on the spread of SARS-CoV-2 variants in the population.

Sequencing of wastewater samples is however currently mainly used to reconstruct the consensus genome sequence of the most prevalent SARS-CoV-2 strain in the sample and LFV are often not investigated. This consensus sequence can be useful to demonstrate that the detected strain in wastewater corresponds to the dominant strain that circulates in individuals within the same community [32]. However, in contrast to clinical samples, only limited quality criteria are in place when sequencing wastewater samples and those available often only apply for consensus sequence construction. The EU recommends the generation of one million reads per sample and a read length of more than 100 bp [7]. A few studies evaluated LFV in wastewater samples, by using local haplotype reconstruction with ShoRAH [33] or iVar using a minimum coverage of 50X, Phred score of ≥30 and a minimal allelic frequency (AF) of 10% [34]. However, none of these studies evaluated their approach on well-defined populations nor determined detection thresholds for retaining LFV. Since multiple VOCs may co-circulate in a given population, their relative abundance is expected to vary and potentially be very low in wastewater samples. While genome consensus variant calling workflows can only identify mutations present at high AFs, LFV calling methods have been specifically designed to call mutations at lower-than-consensus AFs, and are required to detect VOCs in wastewater samples that are present at an AF below 50%. Appropriate tools and statistical approaches should be provided to ensure reliable and comparable collection and analysis of data, because the detection of LFV is challenging due to the drop in confidence of called mutations at low AFs and sequencing coverages [35–37]. High-quality sequencing reads are required to ensure that single nucleotide variants (SNVs) and indels can be reliably called and quantified. Most LFV calling algorithms therefore consider multiple sequencing characteristics such as strand bias, base quality, mapping quality, sequence context and AF [38] to delineate true variants from sequencing errors. Although the viral diversity in multiple WGS-based studies has been explored using several variant calling methods [39–41], they are often not benchmarked against defined viral populations, rendering the feasibility of using these methods for detecting SARS-CoV-2 VOCs in mixed samples for wastewater surveillance largely unknown.

In this study, we evaluate the performance of LFV detection based on targeted SARS-CoV-2 sequencing to detect and quantify mutations present at low abundances. This approach mimics wastewater deep sequencing by means of the Illumina technology. We used mutations that define the B.1.1.7 lineage as a proof-of-concept. Using two real sequencing datasets that were *in silico* modified by either introducing mutations of interest into raw wild-type sequencing datasets or mixing wild-type and mutant raw sequencing data, we provide guidelines for minimum sequencing coverages to detect clade-defining mutations at specific AFs.

## 2 Methods

### 2.1 Employed sequencing data and generation of consensus genome sequences

SARS-CoV-2 raw sequencing data from 316 samples was downloaded from the Sequence Read Archive (SRA) [42]. A random selection of samples was done on the 27^th^ of January 2021 from the COVID-19 Genomics UK (COG-UK) consortium (PRJEB37886) including only samples with a submission date in January 2021, sequenced with Illumina Novaseq 6000 and using an amplicon-based enrichment strategy (Supplementary File S1).

To ensure correct pairing of fastq files, all samples were re-paired using BBMap v38.89 repair.sh with default settings [43] (Figure 1: Step 1). The consensus genome sequences were generated for all these samples (Figure 1: Step 2). The workflow was built using the Snakemake workflow management system using python 3.6.9 [44]. Next, the re-paired paired-end reads were trimmed using Trimmomatic v0.38 [45] setting the following options: ‘LEADING:10’, ‘TRAILING:10’ ‘SLIDINGWINDOW:4:20’, and ‘MINLEN:40’. As reference genome, the sequence with GISAID [25] accession number EPI_ISL_837246 was used for the wild-type samples, while EPI_ISL_747518 was used for the mutant samples. These reference genomes were indexed using Bowtie2-build v2.3.4.3 [46]. Trimmed reads were aligned to their respective reference genomes using Bowtie2 v2.3.4.3 [46] using default parameters. The resulting SAM files were converted to BAM files using Samtools view v1.9 [47] and sorted and indexed using the default settings of respectively Samtools sort and Samtools index v1.9 [47]. Using the sorted BAM file, a pileup file was generated with Samtools mpileup v1.9 [47] using the options “--count-orphans” and “--VCF”. Next, the variants were called with bcftools call v1.9 [47] using the options “-O z”, “--consensus-caller”, “--variants-only” and “ploidy 1”, and converted and indexed to uncompressed VCF files with respectively bcftools view v1.9 [47] using the options “--output-type v” and bcftools index v1.9 [47] using the option “--force”. Lastly, a temporary consensus sequence was generated using bcftools consensus v1.9 [47] with default settings, providing the reference genome and produced VCF file as inputs. Afterwards, the previous steps were repeated once with the same options using the generated temporary consensus sequence as fasta reference to generate the final consensus sequence. These sequences were used to confirm either the presence or absence of the clade-defining mutations of the B.1.1.7 mutant for both the mutant and wild-type samples respectively (Table 1). To extract the sequencing coverage for each position and subsequently calculate the median coverage for each sample, Samtools depth v1.9 [47] was used on the BAM files. Additionally, bamreadcount v0.8.0 (https://github.com/genome/bam-readcount) was run on all samples using the BAM files to determine the coverage at each position.

**Table 1:**
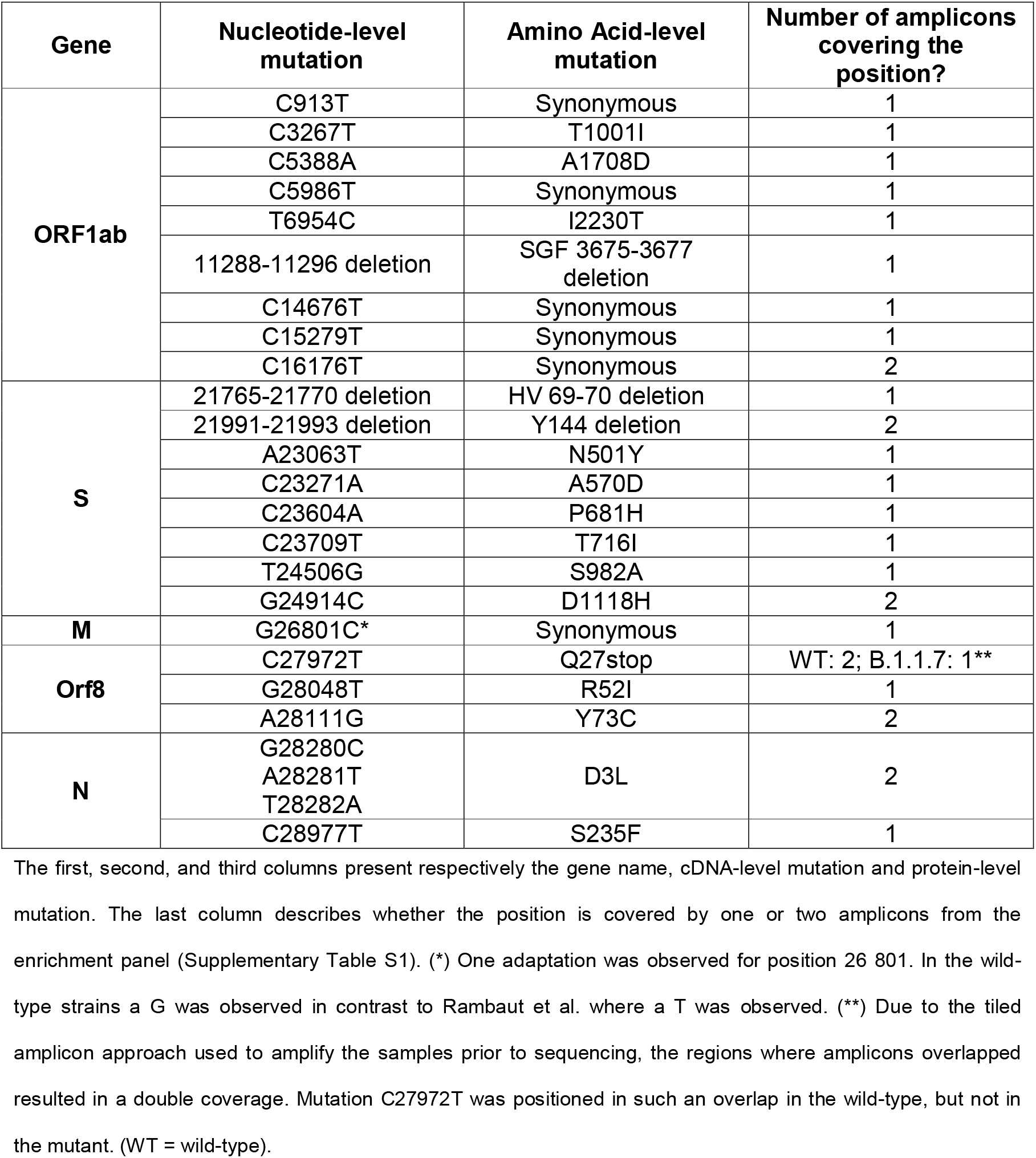
Mutations linked to SARS-CoV-2 lineage B.1.1.7 [48].

**Figure 1:**
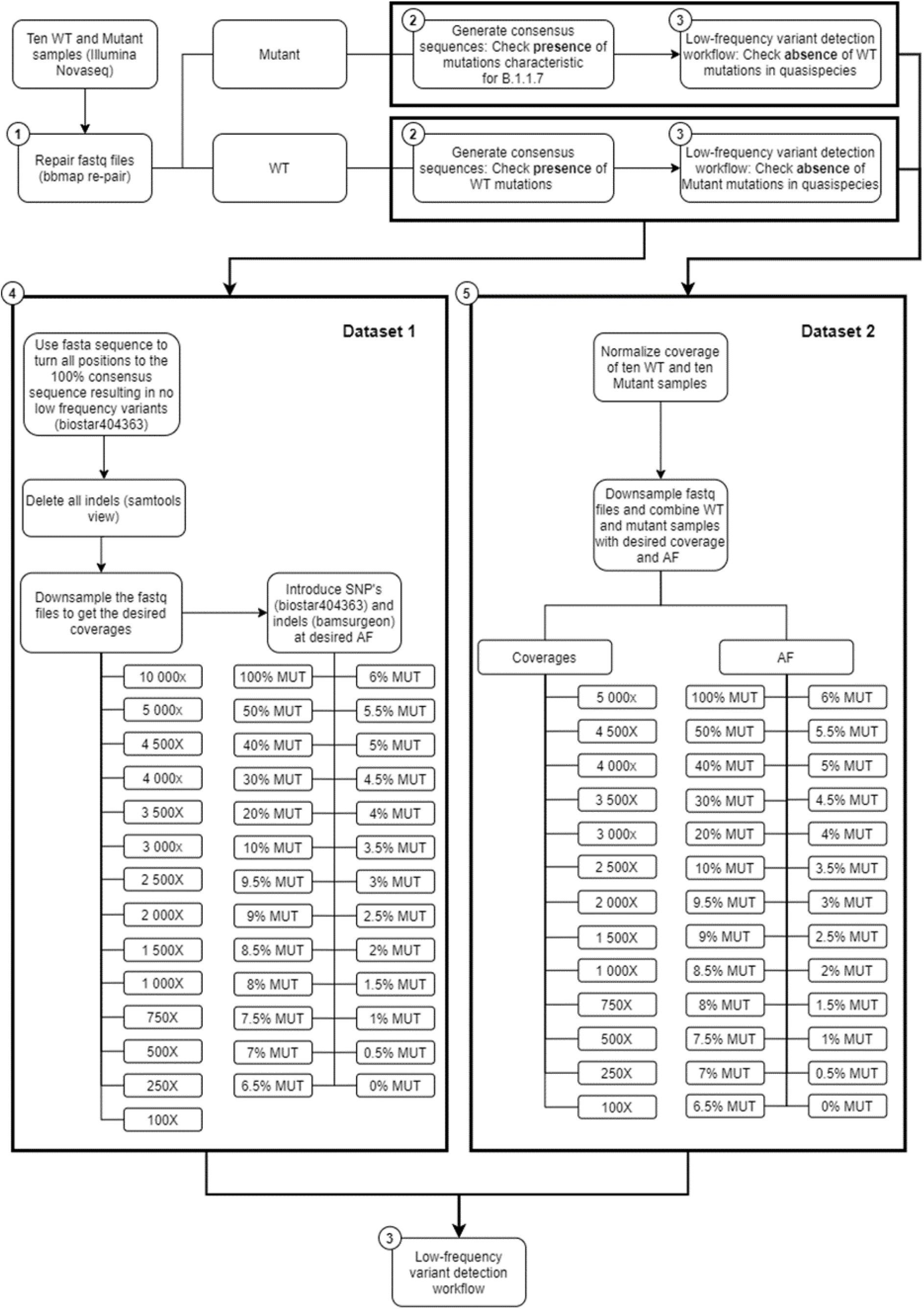
Schematic representation of the workflow.

From the initial 316 samples, ten mutant samples were selected that presented similar coverage depth at the positions of interest after normalization (see below). These samples contained the mutations assigned to the B.1.1.7 variant. Ten wild-type samples were also chosen that did not contain any of these mutations (Table 1, Table 2) and also presented similar coverage depth at the positions of interest after normalization. Lineage B.1.1.7, termed Variant of Concern (VOC) 202012/01 by Public Health England (PHE) [49], 20I/501Y.V1 by Nextstrain [50] and alpha variant by the World Health Organisation [51], was first reported in the United Kingdom but its prevalence continues to rise in many European countries [52]. This variant was found to be more transmissible [17] and may cause more severe infections [18, 19]. Lineage B.1.1.7 is defined by multiple spike protein changes, including deletion 69-70 and deletion 144 in the N-terminal domain, amino changes N501Y in the receptor-binding domain, and amino acid changes A570D, P681H, T716I, S982A, D1118H, as well as mutations in other genomic regions [53]. More recently PHE has reported B.1.1.7 cases with an additional mutation, E484K [49]. Median coverages of the selected samples were consistently high (minimum 13,848; maximum 36,255) and median read lengths were always 221 and 201 for the forward and reverse reads respectively (Table 2). Additionally, as suggested by ECDC, more than 95% of the genome was covered by reads with a minimal coverage of 500X [31].

**Table 2:**
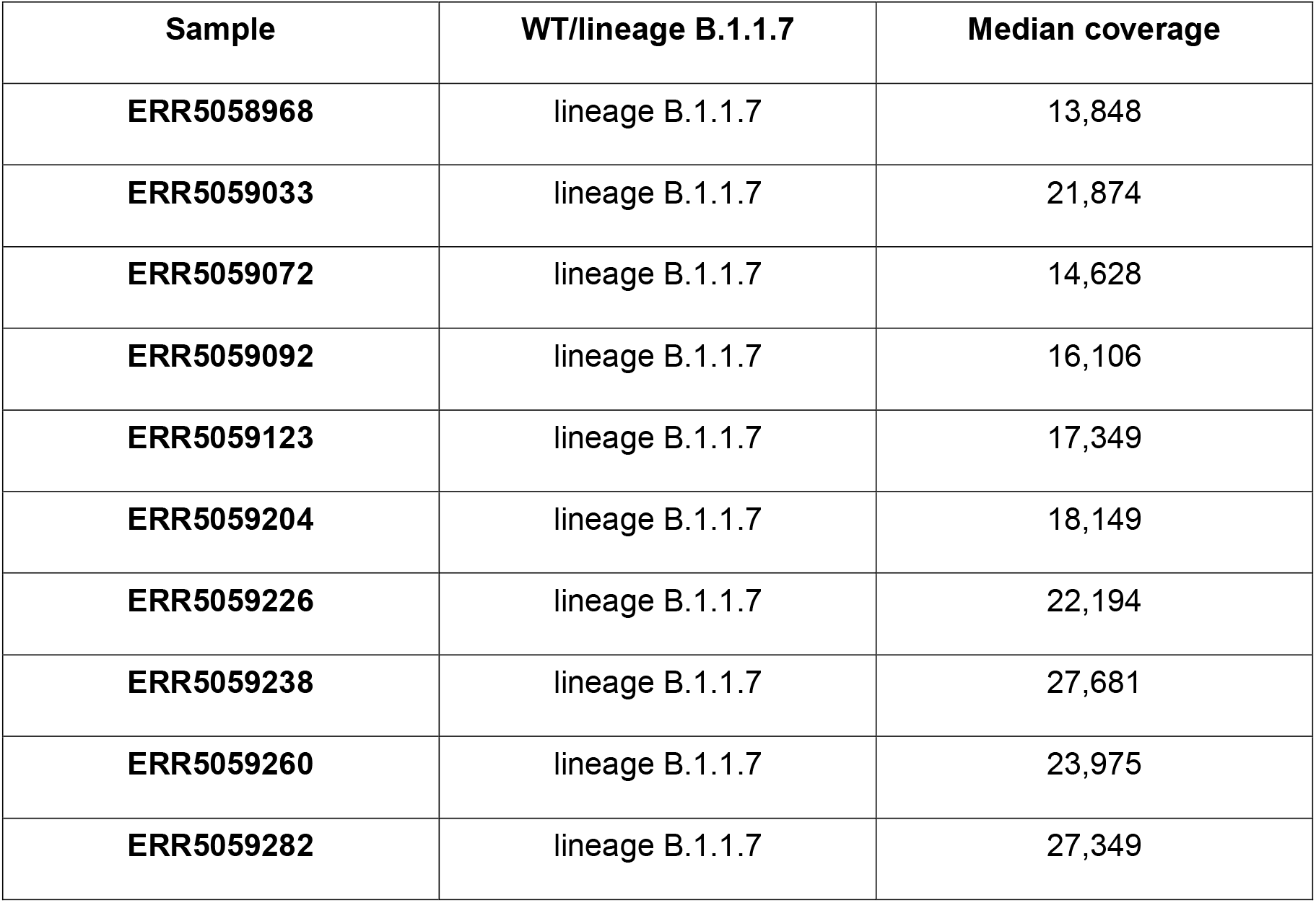
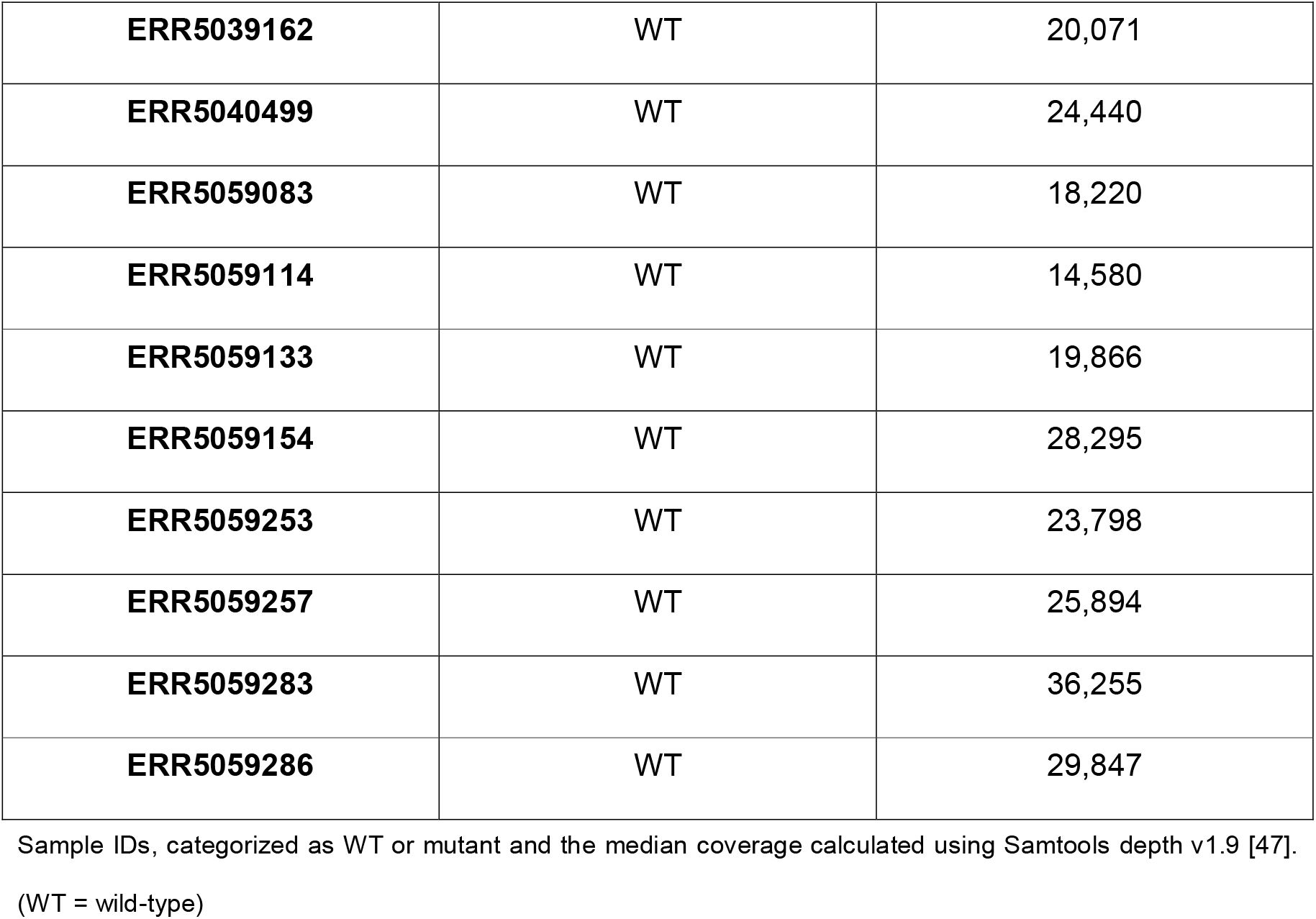
List of SRA accession numbers used for employed wild-type and lineage B.1.1.7 samples in this study.

| Sample | WT/lineage B.1.1.7 | Median coverage |
| --- | --- | --- |
| ERR5058968 | lineage B.1.1.7 | 13,848 |
| ERR5059033 | lineage B.1.1.7 | 21,874 |
| ERR5059072 | lineage B.1.1.7 | 14,628 |
| ERR5059092 | lineage B.1.1.7 | 16,106 |
| ERR5059123 | lineage B.1.1.7 | 17,349 |
| ERR5059204 | lineage B.1.1.7 | 18,149 |
| ERR5059226 | lineage B.1.1.7 | 22,194 |
| ERR5059238 | lineage B.1.1.7 | 27,681 |
| ERR5059260 | lineage B.1.1.7 | 23,975 |
| ERR5059282 | lineage B.1.1.7 | 27,349 |
| **ERR5039162** | WT | 20,071 |
| **ERR5040499** | WT | 24,440 |
| **ERR5059083** | WT | 18,220 |
| **ERR5059114** | WT | 14,580 |
| **ERR5059133** | WT | 19,866 |
| **ERR5059154** | WT | 28,295 |
| **ERR5059253** | WT | 23,798 |
| **ERR5059257** | WT | 25,894 |
| **ERR5059283** | WT | 36,255 |
| **ERR5059286** | WT | 29,847 |

Sample IDs, categorized as WT or mutant and the median coverage calculated using Samtools depth v1.9 [47].
(WT = wild-type)

### 2.2 LFV detection

The absence of pre-existing wild-type and mutant LFV at the positions defining lineage B.1.1.7 (Table 1) was verified in both the mutant and wild-type samples (Figure 1: Step 3), respectively, by calling all LFV in these samples and subsequently checking the positions of interest. Python 3.6.9 was used with the packages pysam 0.16.0.1 [54] and numpy 1.19.5 [55]. Each generated (final) consensus FASTA file was used as reference for its respective sample and indexed using Samtools faidx v1.9 [47] and Bowtie2-build v2.3.4.3. Bowtie2 v2.3.4.3 was then used to align the reads of each sample to its reference sequence, producing a SAM file that was converted into BAM using Samtools view v1.9. Next, reads were sorted using Picard SortSam v2.18.14 (https://github.com/broadinstitute/picard) with the option “SORT_ORDER=coordinate” and Picard CreateSequenceDictionary v2.18.14 [56] was used to generate a dictionary of the reference FASTA file. Picard AddOrReplaceReadGroups v2.18.14 [56] was afterwards run on the reads with the flags “LB”, “PL”, “PU” and “SM” set to the arbitrary placeholder value “test”. The resulting BAM files were indexed using Samtools index v1.9 and used as input for GATK RealignerTargetCreator 3.7 [57], which was followed by indel realignment using GATK IndelRealigner v3.7 [57]. Next, generated BAM files were indexed using Samtools index v1.9. The call function of the LoFreq v2.1.3.1 package [36] was used to call LFV in the BAM files and generate a VCF file using the options “--call-indels” and “--no-default-filter” and using the consensus sequence as reference to call LFV. Next, the unfiltered VCF file was filtered using the filter function of the LoFreq v2.1.3.1 package, setting the strand bias threshold for reporting a variant to the maximum allowed value by using the option “--sb-thresh 2147483647” to allow highly strand-biased variants to be retained, to account for the non-random distribution of reads due to the design of the amplification panel. All employed scripts are available in Supplementary File S2. Additionally, the workflow is also available at the public Galaxy instance of our institute at https://galaxy.sciensano.be as a free resource for academic and non-profit usage. The presence of the nucleotides assigned to the B.1.1.7 lineage or the wild-type (Table 1) was verified for the mutant and wild-type samples, respectively. Additionally, it was checked that there were no LFV at these positions, so that the wild-type nucleotide or mutant nucleotide was always present at 100% for the retained 10 WT and 10 mutant samples.

#### 2.2.1 Dataset 1: *In silico* insertion of mutations of interest into raw sequencing datasets

For the first dataset (Figure 1: Step 4), all low-frequency single nucleotide polymorphisms (SNPs) were removed from the raw sequencing data of all samples. SNPs were removed using Jvarkit employing biostar404363 [58] by converting all nucleotides to the consensus fasta sequence. Next, all ten WT samples were down-sampled using “seqtk sample” with argument “-s100” (https://github.com/lh3/seqtk) to 14 different (median) coverages (100X, 250X, 500X, 750X, 1000X, 1500X, 2000X, 2500X, 3000X, 3500X, 4000X, 4500X, 5000X and 10,000X). The 22 SNP mutations characteristic for the B.1.1.7 lineage (Table 1) were introduced at 26 different AF (mutant: 0%, 0.5%, 1%, 1.5%, 2%, 2.5%, 3%, 3.5%, 4%, 4.5%, 5%, 5.5%, 6%, 6.5%, 7%, 7.5%, 8%, 8.5%, 9%, 9.5%, 10%, 20%, 30%, 40%, 50%, 100%) at the various coverages mentioned above employing biostar404363. This resulted in 10 samples at 364 conditions (i.e. combination of coverage and AF). Next, all reads containing indels were removed from these samples using samtools view v1.9. Finally, the three deletions associated with the B.1.1.7 lineage were introduced at the 26 AF mentioned above using BAMSurgeon 1.2 [59], which was adapted to decrease runtime, with the options “-p 10”, “--force”, “-d 0”, “--ignorepileup”, “--mindepth 1”, “--minmutreads 1”, “--maxdepth 1000000”, “--aligner mem”, “--tagreads”. A minority of reads that were lacking a mate in the targeted regions were removed by using an in-house script making use of Python 3.6.9 and the package pysam 0.16.0.1. Samples in BAM format were then converted back to FASTQ format using bedtools bamtofastq v2.27.1 [60]. Finally the LFV detection workflow (Figure 1: Step 3) described in section 2.2 was used on these 10 samples for all 364 conditions using the FASTA file of the wild-type sample as reference with LoFreq.

#### 2.2.2 Dataset 2: Introduction of mutations of interest by mixing wild-type and mutant raw sequencing read datasets

For the second dataset (Figure 1: Step 5), the coverage of all 20 samples (Table 2) was normalized to 5000X using BBMap v38.89 bbnorm.sh [43] with the options “target=5000”, “mindepth=5”, “fixspikes=f”, “passes=3”, “uselowerdepth=t”. However, due to the tiled amplicon approach used to amplify these samples prior to sequencing, regions where amplicons overlapped subsequently had double coverage resulting in two coverages, i.e. 5000X and 10,000X, after normalization (Supplementary Table S1). *In silico* datasets were then generated by mixing the appropriate number of reads for every combination of the ten wild-type and ten mutant samples, resulting in a total of 100 mixed samples, which were down-sampled using “seqtk sample” (with option “–s100”) to the appropriate fractions for the required combination of 13 final coverages (100X, 250X, 500X, 750X, 1000X, 1500X, 2000X, 2500X, 3000X, 3500X, 4000X, 4500X and 5000X) and 26 AF (mutant: 0%, 0.5%, 1%, 1.5%, 2%, 2.5%, 3%, 3.5%, 4%, 4.5%, 5%, 5.5%, 6%, 6.5%, 7%, 7.5%, 8%, 8.5%, 9%, 9.5%, 10%, 20%, 30%, 40%, 50%, 100%). This resulted in 100 mixed samples at 338 conditions (i.e. combination of coverage and AF). Finally, the LFV detection workflow (Figure 1: Step 3) described in section 2.2 was used on these samples for all conditions using the FASTA file of the wild-type sample as reference, except for samples mimicking 100% AF for the mutant positions where the FASTA file of the mutant sample was used.

Although the second dataset was normalized for total coverage at every genomic position, the tiled amplicon approach resulted in some genomic positions being covered by two overlapping amplicons. Two groups of mutations were therefore obtained for every coverage (Table 2), i.e. for a targeted coverage of 5000X, 17 mutations were present at ∼5000X (C913T, C3267T, C5388A, C5986T, T6954C, 11288-11296 deletion, C14676T, C15279T, 21765-21770 deletion, A23063T, C23271A, C23604A, C23709T, T24056G, G26801C, G28048T, C28977T) and 7 mutations were present at ∼10,000X (T16176C, 21991-21993 deletion, G24914C, A28111G, G28280C, A28281T, T28282A). Mutation C27972T was excluded from further analysis, because this position in the wild-type samples was located in a region where amplicons overlapped resulting in a coverage of approximately 10,000X, while in mutant samples it was in a region with no overlap and where a coverage of 5000X was therefore observed (Supplementary Table S1). For further analysis, the results were pooled together per theoretical coverage resulting in 24 mutations per coverage but only 17 and 7 mutations at the lowest (i.e. 100X) and highest (i.e. 10,000X) coverage, respectively (Supplementary Table S2). The actual median coverage was calculated per theoretical targeted coverage using the output of bamreadcount v0.8.0 of each sample. Using this output, the coverage of each position of interest was extracted (Supplementary Table S2).

### 2.3 Qualitative evaluation of detection of B.1.1.7 at different abundances

Since samples of Dataset 1 were normalized for the total median coverage, different individual positions of interest could exhibit deviating coverages. For the qualitative evaluation of LFV detection (i.e. can mutant positions of interest be correctly detected?), the number of false negatives were counted per condition (i.e. combination of AF and coverage) and divided by the total number of observations (i.e. the number of samples (n=10) and number of mutations considered for that condition (n=25)). A mutant position of interest was considered as correctly detected as soon as it was detected by LoFreq, irrespective of its estimated AF.

Dataset 2 was subjected to the same qualitative evaluation as described for Dataset 1. The number of false negatives per condition was divided by the number of observations (i.e. the number of samples (n=100) and number of mutations considered for that condition (either n=7, n=17 or n=24)).

The visualisation of the qualitative evaluation was performed using a contour plot from the R package plotly (RStudio 1.0.153; R3.6.1) [61]. The false negative (FN) proportion in the qualitative evaluation plots ranged from 0 to 1 with a step size of 0.1.

### 2.4 Quantitative evaluation of detection of B.1.1.7 at different abundances

For the quantitative evaluation of LFV detection (i.e. is the estimated AF of correctly detected mutant positions of interest close to the true AF?) of both datasets, FN values were considered as ‘below the quantification limit’ with the quantification limit equal to the lowest recorded value for that condition (i.e. combination of AF and coverage). Outliers were identified for each condition using the Grubbs test that was sequentially applied by first searching for two outliers at the same side, followed by a search for exactly one outlier. If the p-value of the Grubbs test was below 0.05, outliers were excluded. The standard deviation (SD) and mean value of AF for every condition were estimated by a maximum likelihood model based on the normal distribution that took the FN into account as censor data. Data were modelled according to a normal distribution. If the percentage of FN results was above 75%, the condition was however excluded from quantitative evaluation. Finally, a performance metric describing closeness to the true AF was calculated for each targeted AF individually by dividing each pooled squared SD by the maximal pooled squared SD. This metric will range between 0, relatively the closest to the targeted AF, and 1, relatively the furthest from the targeted AF.

As described for the qualitative evaluation, contour plots from the R package plotly (RStudio 1.0.153; R3.6.1) were used for the visualisation of the quantitative evaluation. The performance metric in the quantitative evaluation plots ranged from 0 to 1 with a step size of 0.1.

## 3 Results

### 3.1 Qualitative evaluation demonstrates that B.1.1.7 clade-defining mutations can be reliably detected at low AF when sequencing coverage is adequately high

To mimic targeted SARS-CoV-2 sequencing with a VOC present at low abundances in the viral population, B.1.1.7 clade-defining mutations were first *in silico* introduced at well-defined AFs and coverages in real sequencing data (‘Dataset 1’) of ten wild-type samples, without however using any coverage normalization so that individual mutations could be present at higher or lower coverages compared to the total median genomic coverage due to unevenness of coverage. To assess whether introduced mutations were correctly detected, or alternatively missed as FN, samples of this dataset were analysed using a LFV calling workflow based on LoFreq.

Figure 2A depicts the proportion of FN observations, and corresponding values are presented in Table 3, for all evaluated coverages and targeted AFs until 20%. Results for all targeted AFs (including higher values) are presented in Supplementary Figure S1 and Supplementary Table S3. All LFV could be detected at an AF of 1% at a median coverage of 10,000X. As the coverage decreased, the AF threshold at which no single FN occurred (i.e. perfect sensitivity) increased to 1.5% at 5000X, 3% at 1000X, 5% at 500X, 9.5% at 250X, and 20% at 100X. When allowing a maximum of 10% FN (i.e. sensitivity of 90%), the AF thresholds decreased substantially to 1% at 5000X, 1.5% at 1000X, 2.5% at 500X, 4% at 250X, and 7.5% at 100X. No false positive mutations related to the mutant and wild-type were observed at respectively 0% and 100% AF.

**Table 3:** Qualitative evaluation of Dataset 1 based on false negative proportions per condition until a targeted mutant AF of 20%. The percentage of FN is coloured ranging from 0 (dark) to 1 (light) according to the gradient depicted in Figure 2A.

| Coverage →<br>AF ↓ | 100 | 250 | 500 | 750 | 1000 | 1500 | 2000 | 2500 | 3000 | 3500 | 4000 | 4500 | 5000 | 10,000 |
| --- | --- | --- | --- | --- | --- | --- | --- | --- | --- | --- | --- | --- | --- | --- |
| 20.00% | 0% | 0% | 0% | 0% | 0% | 0% | 0% | 0% | 0% | 0% | 0% | 0% | 0% | 0% |
| 10.00% | 5% | 0% | 0% | 0% | 0% | 0% | 0% | 0% | 0% | 0% | 0% | 0% | 0% | 0% |
| 9.50% | 4% | 0% | 0% | 0% | 0% | 0% | 0% | 0% | 0% | 0% | 0% | 0% | 0% | 0% |
| 9.00% | 7% | 1% | 0% | 0% | 0% | 0% | 0% | 0% | 0% | 0% | 0% | 0% | 0% | 0% |
| 8.50% | 4% | 0% | 0% | 0% | 0% | 0% | 0% | 0% | 0% | 0% | 0% | 0% | 0% | 0% |
| 8.00% | 9% | 2% | 0% | 0% | 0% | 0% | 0% | 0% | 0% | 0% | 0% | 0% | 0% | 0% |
| 7.50% | 8% | 0% | 0% | 0% | 0% | 0% | 0% | 0% | 0% | 0% | 0% | 0% | 0% | 0% |
| 7.00% | 10% | 1% | 0% | 0% | 0% | 0% | 0% | 0% | 0% | 0% | 0% | 0% | 0% | 0% |
| 6.50% | 15% | 2% | 0% | 0% | 0% | 0% | 0% | 0% | 0% | 0% | 0% | 0% | 0% | 0% |
| 6.00% | 15% | 1% | 0% | 0% | 0% | 0% | 0% | 0% | 0% | 0% | 0% | 0% | 0% | 0% |
| 5.50% | 19% | 3% | 0% | 0% | 0% | 0% | 0% | 0% | 0% | 0% | 0% | 0% | 0% | 0% |
| 5.00% | 22% | 3% | 0% | 0% | 0% | 0% | 0% | 0% | 0% | 0% | 0% | 0% | 0% | 0% |
| 4.50% | 26% | 4% | 2% | 0% | 0% | 0% | 0% | 0% | 0% | 0% | 0% | 0% | 0% | 0% |
| 4.00% | 31% | 6% | 1% | 0% | 0% | 0% | 0% | 0% | 0% | 0% | 0% | 0% | 0% | 0% |
| 3.50% | 45% | 12% | 4% | 0% | 0% | 0% | 0% | 0% | 0% | 0% | 0% | 0% | 0% | 0% |
| 3.00% | 47% | 18% | 4% | 1% | 0% | 0% | 0% | 0% | 0% | 0% | 0% | 0% | 0% | 0% |
| 2.50% | 62% | 21% | 7% | 2% | 2% | 0% | 0% | 0% | 0% | 0% | 0% | 0% | 0% | 0% |
| 2.00% | 70% | 32% | 14% | 7% | 3% | 0% | 0% | 0% | 0% | 0% | 0% | 0% | 0% | 0% |
| 1.50% | 84% | 52% | 24% | 16% | 9% | 5% | 1% | 2% | 0% | 0% | 0% | 0% | 0% | 0% |
| 1.00% | 96% | 77% | 54% | 35% | 28% | 15% | 8% | 6% | 6% | 3% | 2% | 2% | 2% | 0% |
| 0.50% | 98% | 95% | 85% | 77% | 70% | 57% | 46% | 41% | 33% | 29% | 22% | 22% | 16% | 7% |
| 0.00% | 0% | 0% | 0% | 0% | 0% | 0% | 0% | 0% | 0% | 0% | 0% | 0% | 0% | 0% |

**Figure 2:**
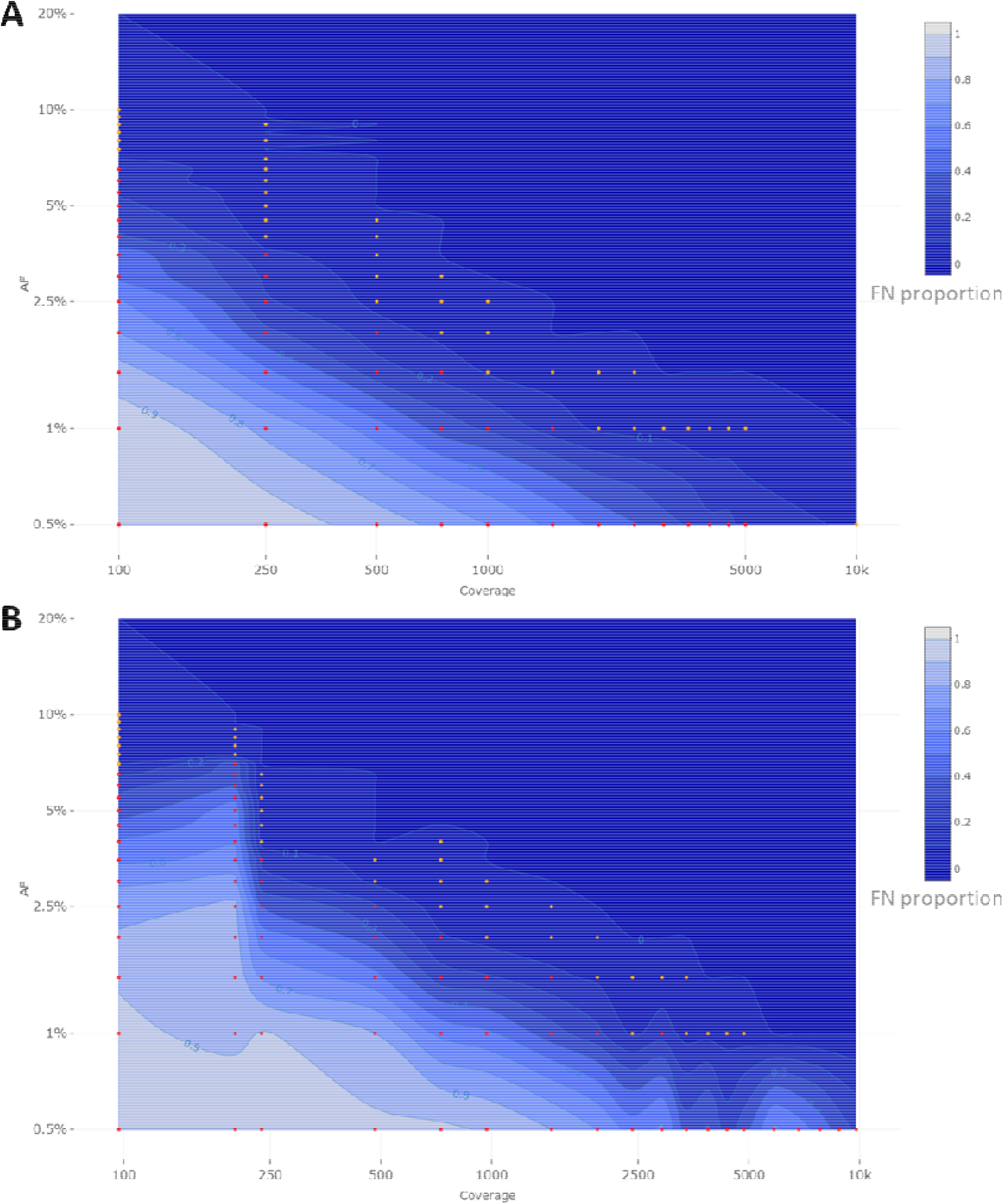
Qualitative evaluation of Dataset 1 (A) and Dataset 2 (B) based on false negative proportions per condition until a targeted mutant AF of 20%. Orange and red dots represent conditions with a FN proportion between 0 and 0.1, and between 0.1 and 1, respectively. The percentage of FN is coloured ranging from 0 (dark) to 1 (light) in intervals of 0.1 as extrapolated using a contour plot in the R package plotly [61] (actual FN proportions are presented in Table 3 for Dataset 1 and Table 4 for Dataset 2). Results for targeted mutant AF values >20% are presented in Supplementary Figure S1 for Dataset 1 and Supplementary Figure S2 for Dataset 2. Both the x- and y-axis follow a logarithmic scale.

A second approach was also considered for mimicking targeted SARS-CoV-2 virus sequencing with a VOC present at low abundances, by *in silico* mixing real raw sequencing reads from ten B.1.1.7 samples into ten wild-type samples (‘Dataset 2’) for a total of 100 mixes at well-defined AFs and coverages, while applying coverage normalization so that individual mutations were present at approximately similar coverages for all B.1.1.7 clade-defining positions.

**Table 4:**
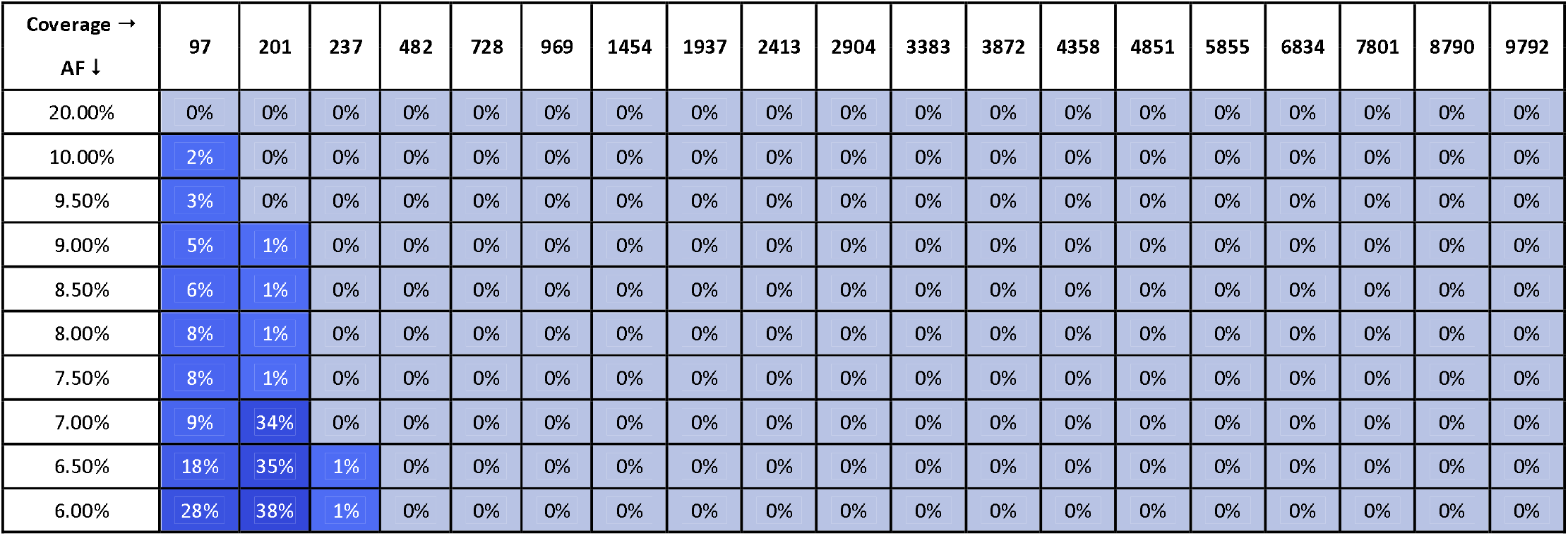
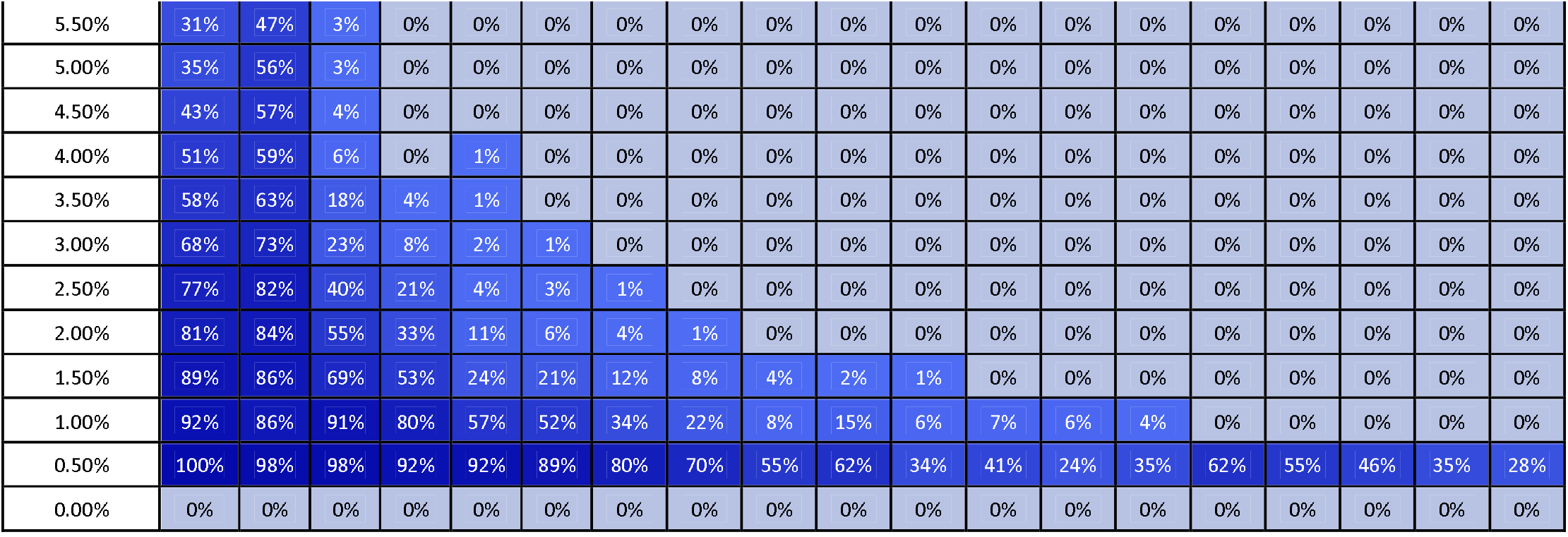
Qualitative evaluation of Dataset 2 based on false negative proportions per condition until a targeted mutant AF of 20%. The percentage of FN is coloured ranging from 0 (dark) to 1 (light) according to the gradient depicted in Figure 2B.

Figure 2B depicts the proportion of FN observations, and actual values are presented in Table 4, for all evaluated coverages and targeted AF until 20%. Results for higher targeted AF are presented in Supplementary Figure S2 and Supplementary Table S4. All LFV could be detected at an AF of 1% at a median coverage of 9792X. As the coverage decreased, the AF thresholds at which no single FN occurred (i.e. perfect sensitivity) increased to 1.5% at 4851X, 3.5% at 969X, 4% at 482X, 7% at 237X, and 20% at 97X. However, when allowing a maximum of 10% FN (i.e. reducing the sensitivity to 90%), the AF thresholds decreased substantially to 1% at 4851X, 2% at 969X, 3% at 482X, 4% at 237X, and 7% at 97X. No false positive mutations related to the mutant and wild-type were observed at 0% and 100%. Overall, the results for Dataset 1, using the median coverages, and Dataset 2, using the coverages at the positions of interest, were qualitatively similar.

### 3.2 Quantitative evaluation demonstrates that the resulting AFs for B.1.1.7 clade-defining mutations are close to their target values

To evaluate the possibility of quantifying LFV in both datasets, the SDs of available observations were first evaluated for each condition (i.e. combination of AF and coverage). This provisional analysis indicated that for both Dataset 1 (Supplementary File S3) and Dataset 2 (Supplementary File S4), the SD systematically decreased per target AF as coverage increased. This provisional analysis also indicated that for both datasets, irrespective of coverage, the SD generally increased between a targeted AF of 1% to 10%, after which it plateaued for targeted AFs above 20%. We therefore employed the squared SD per AF divided by the maximal squared SD per target AF to describe closeness of observed AF to the true AF, for which results are presented in Figure **3**A for Dataset 1. As expected, the variation in AF estimates fluctuates in function of the median coverage and targeted AF, with variation decreasing per target AF as coverage increased, but also variation being generally more pronounced at low AFs irrespective of coverage. Notwithstanding, even for regions in Figure 3A exhibiting high variation, the variability overall remained small (Supplementary File S3). The interquartile range (IQR) (Supplementary File S3D) of the observed AF was still limited at the various targeted AF ranging from 0.62%-6.26% at an AF of 50%, 0.36%-3.49% at an AF of 10% and 0.27%-2.07% at an AF of 5% with the highest IQR observed at lower coverages.

**Figure 3:**
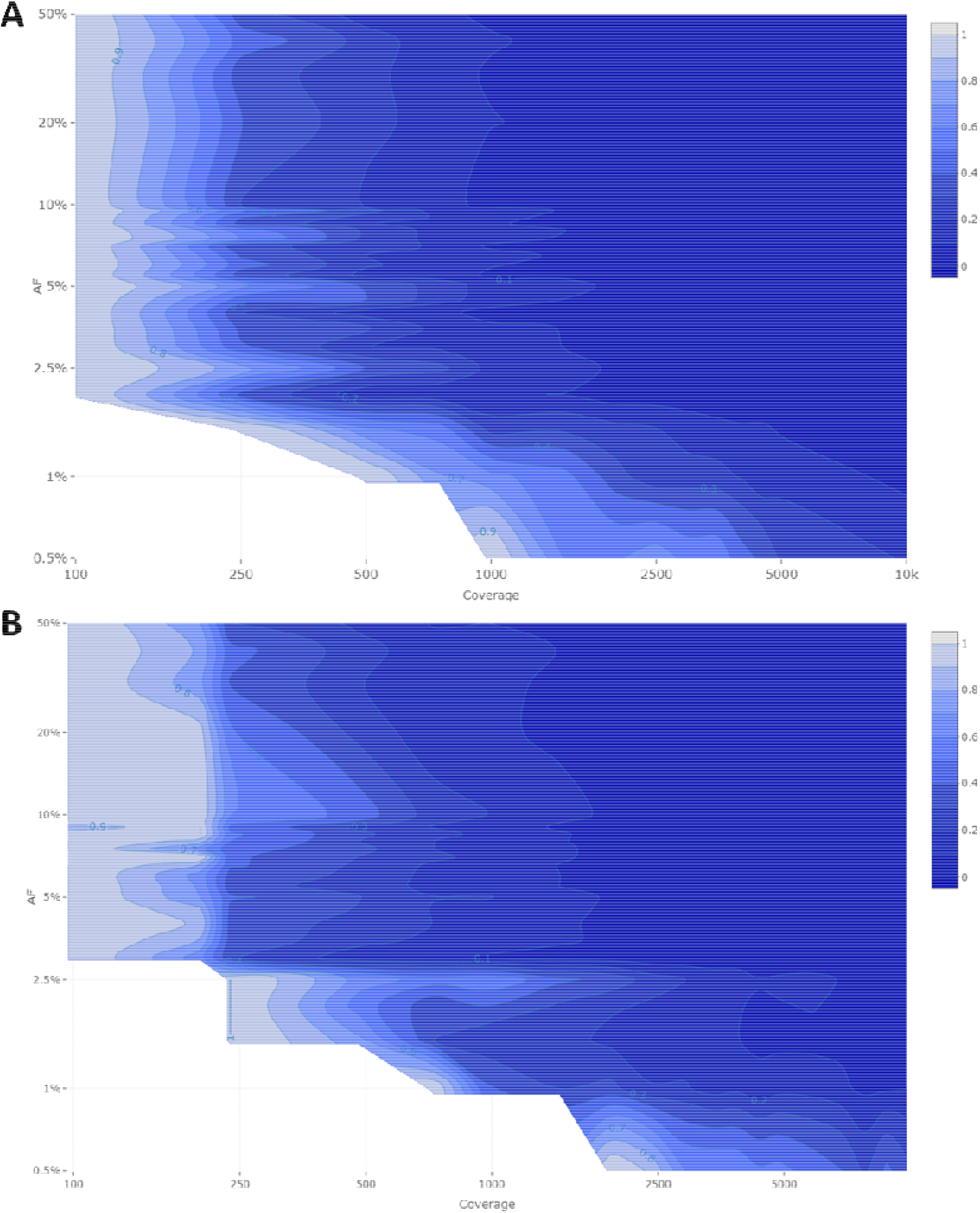
Quantitative evaluation of Dataset 1 (A) and Dataset 2 (B) using the squared SD divided by the maximal squared SD per targeted AF. The figure is coloured ranging from 0 (dark) to 1 (light) in intervals of 0.1 as extrapolated using a contour plot in the R package plotly [61] (actual values are presented in Supplementary File S3 for Dataset 1 and Supplementary File S4 for Dataset 2). Both the x- and y-axis follow a logarithmic scale. Conditions with a FN proportion higher than 75% were excluded and correspond to the white plane in the lower left corner.

Results for the quantitative evaluation of Dataset 2 are presented in Figure 3B, and are in accordance with the trends observed for Dataset 1 with the variation decreasing per target AF as coverage increased, and lower target AFs exhibiting increasing variation irrespective of coverage. Notwithstanding, similarly to Dataset 1, the observed total variation remained small (Supplementary File S4). The IQR (Supplementary File S4D) of the observed AF was limited at the various targeted AF ranging from 0.73%-3.93% at an AF of 50%, 0.41%-3.93% at an AF of 10% and 0.29%-2.27% at an AF of 5% with the highest IQR observed at lower coverages.

## 4 Discussion

Wastewater surveillance has been recommended to be used in the EU for improving the epidemiological surveillance of SARS-CoV-2 [7]. WGS is a more suitable approach than RT-qPCR to track both existing and newly emerging SARS-CoV-2 variants. Wastewater sequencing is currently however mainly used to construct the consensus genome sequence and determine the most prevalent strain in communities, but interest exists in its potential for detecting LFV and consequently determining all circulating variants, in particular VOCs [7].

To evaluate the potential of targeted amplicon-based SARS-CoV-2 WGS to detect and quantify VOCs present at low abundances in mixed samples, we assessed the performance of a workflow designed for LFV detection in WGS data of wastewater samples. Mutations defining lineage B.1.1.7 were employed as a proof-of-concept using an approach based on *in silico* modifying real sequencing data to construct two datasets, mimicking wastewater deep sequencing with the Illumina technology. For the first dataset, lineage B.1.1.7-defining mutations were introduced *in silico* into raw wild-type sequencing datasets. For the second dataset, the same mutations were introduced by mixing wild-type and B.1.1.7 raw sequencing datasets. In Dataset 1, the coverage profile of the samples corresponded to a typical real dataset including large fluctuations in sequencing coverage at certain positions. In Dataset 2, sequencing coverages were normalized, which allowed evaluating with high precision how reliable AF detection is at specific coverages. Afterwards, the ability to both detect and quantify LFV was evaluated. Results demonstrated that WGS enabled detecting LFV with very high performance. As expected, lower coverages and AFs resulted in lower sensitivity and higher variability of estimated AFs. We found, employing the most conservative thresholds from either Datasets 1 or 2, that a sequencing coverage of 250X, 500X, 1500X, and 10,000X is required to detect all LFV at an AF of 10%, 5%, 3% and 1%, respectively (Table 3 and Table 4). For quantification of variants, the variability remained overall small for all conditions respecting the above thresholds, resulting in reliable abundance estimations, despite the variability of estimated AF increasing at lower coverages and AF. Of note, it was observed that the profile of the genome coverage differed at some positions between wild-type and mutant samples indicating that the amplicon-based enrichment approach could possibly introduce a bias. Consequently, this should be considered when examining and quantifying the proportion of mutants in the sample.

Obtaining high coverages for wastewater samples may however be challenging under real-world conditions. In contrast to clinical samples in which viral loads are typically high, ranging from 10^4^ to 10^7^ copies/mL [62], viral RNA loads in wastewater samples are often low, ranging from 10^−1^ to 10^3.5^ copies/mL [63]. This renders it more challenging to sequence samples with a low viral load. Additionally, variants circulating at low frequencies in a community are expected to be present at a low AF in wastewater samples. Nevertheless, employing the most conservative thresholds from either Datasets 1 or 2, 90% of LFV present at an AF of 10%, 5%, 3% and 1% were still detected at a sequencing coverage of 100X, 250X, 500X, and 2500X respectively (Table 3 and Table 4). This study focussed on the sensitivity of LFV detection and did not explore the false positive rates (i.e. specificity). Although our recommendations for AFs and coverages ensure high sensitivity, often an inverse relationship exists between sensitivity and specificity and we can therefore not exclude that false positives occur for AF and coverage combinations considered as providing qualitative results in this study. A false positive detection is however typically less problematic compared to a false negative result as the former can still be discovered in follow-up investigation in contrast to the latter. Additionally, false positive observations typically occur randomly over the genome [38] and it is unlikely that all VOC-defining mutations would be simultaneously falsely detected, even at low AFs and coverages. The issue of low viral load, low expected AF and potential false positives can be mitigated by sequencing wastewater samples in duplicate when necessary. Possible false positive results could be investigated using RT-qPCR or RT-ddPCR assays that target that specific positions.

Our results can serve as a reference for the scientific community to select appropriate thresholds for the AF and coverage. These could also be context-specific as a smaller or larger degree of false negatives might be warranted for specific applications, and can also be used as a baseline for determining the number of samples that can be multiplexed per run to optimize cost-efficiency of WGS. Our findings highlight the feasibility of using targeted amplicon-based metagenomics approaches for wastewater surveillance, as such samples comprise a collection of different strains, among which the dominant strain will define the consensus sequence of the sample and the detected LFV will represent the circulating strains present at lower frequencies. Other studies that investigated LFV in wastewater provided limited quality criteria regarding the coverage and AF. Furthermore, the quality criteria in these studies were not evaluated using a defined population [33, 34]. ECDC has provided limited quality criteria regarding the sequencing coverage, namely 500X across 95% of the genome to detect LFV, but has not indicated the corresponding AF thresholds this corresponds to for reliable LFV detection [31]. Based on the results obtained in this study, a coverage of 500X allowed to detect LFV until an AF of 5% with perfect sensitivity and would therefore be less suited to detect LFV at lower AFs. Lythgoe et al. recommended a depth of at least 100 reads with an AF of at least 3% to detect the LFV in clinical samples with high viral loads (50,000 uniquely mapped reads) [64]. Based on the results in this study, these recommendations appear not sufficiently strict, since we observed that an AF of 3% requires at least a sequencing coverage of 1500X to detect all LFV or 500X to detect 90% of LFV.

In the presence of multiple VOCs, the VOCs can be identified by composing all possible combinations of LFV as a conservative strategy, although multiple VOCs in one sample will also make the estimation of the relative abundance of each VOC more complicated. If multiple VOCs with partially overlapping defining mutations would be present in a wastewaters sample, some mutations of interest would consequently be present at different AFs. Haplotyping reconstruction methods could be used in such situations to delineate VOCs. However, most haplotype reconstruction programmes perform poorly under higher levels of diversity, and haplotype populations with rare haplotypes are often not recovered [65]. Although haplotype reconstruction has been described for short reads, Nanopore sequencing might offer a substantial advantage for such cases due to its longer reads, despite their higher error rate, to perform haplotype estimation to delineate actual VOCs.

In conclusion, there exists a pressing need for recommendations for detecting LFV for wastewater surveillance. Although further work is still required to investigate the specificity and possibility to detect VOCs instead of just mutations, including for other existing and employed methodologies such as probe-based capture and/or Nanopore sequencing, this study demonstrates the feasibility of a targeted metagenomics approach for highly sensitive LFV detection with acceptable relative abundance estimations using a tiled-amplicon enrichment based on the Illumina technology. This approach enables the detection of mutations associated with specific VOCs. Our approach could also be used to evaluate the potential occurrence of co-infections with other SARS-CoV-2 variants with different strains in clinical samples. In future work this approach should be evaluated on real wastewater data, as in this study high-quality data from clinical specimens was used and modified *in silico* to mimic wastewater data. In light of the pandemic urgency, and the multiple SARS-CoV-2 wastewater surveillance initiatives that are being established and also being integrated into coordinated overarching coordination and preparedness initiatives such as the recently announced European Health Emergency Preparedness and Response Authority [7], we hope that our results will help establishing guidance and recommendations for wastewater surveillance and other relevant applications.

## Data Availability

All data is publicly available.

## Contributions

<u>Conceptualization:</u> Nancy Roosens, Kevin Vanneste, Xavier Saelens; <u>Project Administration</u>: Nancy Roosens; <u>Data Curation</u>: Laura Van Poelvoorde; <u>Methodology</u>: Laura Van Poelvoorde, Thomas Delcourt, Wim Coucke, Sigrid De Keersmaecker, Nancy Roosens, Kevin Vanneste; <u>Software</u>: Laura Van Poelvoorde, Thomas Delcourt, Wim Coucke; <u>Formal Analysis</u>: Laura Van Poelvoorde, Thomas Delcourt, Wim Coucke; <u>Investigation</u>: Laura Van Poelvoorde; <u>Visualization</u>: Laura Van Poelvoorde; <u>Validation</u>: Laura Van Poelvoorde, Thomas Delcourt; <u>Writing – Original Draft Preparation</u>: Laura Van Poelvoorde, Thomas Delcourt, Nancy Roosens, Kevin Vanneste; <u>Writing – Review & Editing</u>: all authors; <u>Funding Acquisition</u>: Nancy Roosens, Philippe Herman; <u>Supervision</u>: Nancy Roosens, Kevin Vanneste

## Ethical disclaimer

Not applicable.

