## Supplemental Files for "Strategy and performance evaluation of low-frequency variant calling for SARS-CoV-2 in wastewater using targeted deep Illumina sequencing"

### Supplementary files

**Supplementary File S1: List of initial 316 samples for which a consensus sequence was generated**

**Supplementary File S2: Scripts for LFV workflow**

#### Lofreq_Workflow.smk

```
from pathlib import Path
import os
root = Path('Path/To/Samples')

all_samples = [fq.name.split('_')[0] for fq in root.iterdir() if '_R1_001.fastq.gz'
in fq.name]

print(all_samples)

snake_dir = workflow.basedir

rule all:
    input:
        CSV = expand(str(root / '{sample}' / 'lofreq' /  '{sample}_nofilter.csv'),
sample=all_samples),

rule trim_reads:
    """
    Trims paired end reads using trimmomatic.
    """
    input:
        FQ_fwd = lambda wildcards: str(root /
f'{wildcards.sample}_R1_001.fastq.gz'),
        FQ_rev = lambda wildcards: str(root /
f'{wildcards.sample}_R2_001.fastq.gz'),
    output:
        FQ_1P = temporary(root / '{sample}' / 'trimming' /
'trimmed_reads_1P.fastq'),
        FQ_2P = temporary(root / '{sample}' / 'trimming' /
'trimmed_reads_2P.fastq'),
        FQ_1U = temporary(root / '{sample}' / 'trimming' /
'trimmed_reads_1U.fastq'),
        FQ_2U = temporary(root / '{sample}' / 'trimming' /
'trimmed_reads_2U.fastq'),
    params:
        basename_output = lambda wildcards: root / wildcards.sample / 'trimming' /
'trimmed_reads.fastq',
        min_len = 40
    threads: 4
    shell:
        """
        module load trimmomatic/0.38;
        cd {params.basename_output.parent}; # Directory is changed because multiple
trimmomatic runs in the same directory interfere with each other
        trimmomatic.sh PE -baseout {params.basename_output} -threads {threads}
{input.FQ_fwd} {input.FQ_rev} \
        ILLUMINACLIP:$TRIMMOMATIC_ADAPTER_DIR/NexteraPE-PE.fa:2:30:10 LEADING:10
TRAILING:10 SLIDINGWINDOW:4:20 \
        MINLEN:{params.min_len}
        """
```

```
rule consensus_fasta_index:
    input:
        FASTA = lambda wildcards: str(root / f"{wildcards.sample.split('-
')[0]}.fasta")
    output:
        INDEX = root / '{sample}' / 'lofreq' / 'reference' / '{sample}.fasta'
    shell:
        """
        module load samtools/1.9;
        module load bowtie2/2.3.4.3;
        cp {input.FASTA} {output.INDEX};
        samtools faidx {output.INDEX};
        bowtie2-build {output.INDEX} {output.INDEX};
        """

rule bt2_map_reads:
    input:
        FASTA = rules.consensus_fasta_index.output.INDEX,
        FQ_1P = rules.trim_reads.output.FQ_1P,
        FQ_2P = rules.trim_reads.output.FQ_2P,
        FQ_1U = rules.trim_reads.output.FQ_1U,
        FQ_2U = rules.trim_reads.output.FQ_2U
    output:
        SAM = temporary(root / '{sample}' / 'lofreq' / '{sample}.sam')
    threads: 16
    shell:
        """
        module load bowtie2/2.3.4.3;
        bowtie2 -x {input.FASTA} -1 {input.FQ_1P} -2 {input.FQ_2P} -S {output.SAM}
-p {threads};
        """

rule sam_to_bam:
    input:
        SAM = rules.bt2_map_reads.output.SAM
    output:
        BAM = temporary(root / '{sample}' / 'lofreq' /  '{sample}.bam')
    shell:
         """
         module load samtools/1.9;
         samtools view -S -b {input.SAM} > {output.BAM};
                  """

rule sort_bam:
    input:
        BAM = rules.sam_to_bam.output.BAM
    output:
        SORT_BAM = temporary(root / '{sample}' / 'lofreq' / '{sample}_sort.bam'),
        SORT_BAM_index = root / '{sample}' / 'lofreq' /  '{sample}_sort.bam.bai'
    shell:
         """
         module load samtools/1.9;
         samtools sort -o {output.SORT_BAM} --output-fmt bam --threads 8
{input.BAM}
         samtools index {output.SORT_BAM};
         """

rule samtools_depth:
    input:
        SORTBAM = rules.sort_bam.output.SORT_BAM
```

```
    output:
        DEPTH = temporary(root / '{sample}' / 'lofreq' / '{sample}_Depth.txt')
    shell:
         """
         module load samtools/1.9;
         samtools depth -d 1000000 {input.SORTBAM} -aa | awk '{{print $1, $3}}' > 
{output.DEPTH}
         """

rule median_average_depth:
    input:
        DEPTH = {rules.samtools_depth.output.DEPTH}
    output:
        AV_DEPTH = root / '{sample}' / 'lofreq' / 
'{sample}_Depth_AverageCoverage.txt',
        MED_DEPTH = root / '{sample}' / 'lofreq' / 
'{sample}_Depth_MedianCoverage.txt'
    params:
        base = lambda wildcards: wildcards.sample
    shell:
         """
         sort -V {input.DEPTH} | awk '{{a[i++]=$2; }} END {{print "{params.base}" 
";" a[int(i/2)]; }}' > {output.MED_DEPTH}
         sort -V {input.DEPTH} | awk '{{total += $2 }} END {{print "{params.base}" 
";" total/NR; }}' > {output.AV_DEPTH}
         """
rule bam_read_count:
    input:
        MED_DEPTH = rules.median_average_depth.output.MED_DEPTH,
        SORTBAM = rules.sort_bam.output.SORT_BAM,
        FASTA = rules.consensus_fasta_index.output.INDEX,
    output:
        TXT = root / '{sample}' / 'lofreq' / '{sample}_bamreadcount.txt'
    params:
        script = 'bam-readcount/bin/bam-readcount',
    resources:
        mem_mb=80000
    shell:
         """
         var=$(grep -e "^>" {input.FASTA} | awk 'sub(/^>/, "")')
         {params.script} -f {input.FASTA} {input.SORTBAM} -w 1 -d 10000000 $var:1-
29861 > {output.TXT}
         """

rule create_seq_dict:
    input:
        TXT = rules.bam_read_count.output.TXT,
        FASTA = rules.consensus_fasta_index.output.INDEX,
    output:
        DICT = root / '{sample}' / 'lofreq' / 'reference' / '{sample}.dict'
    shell:
         """
         module load picard/2.18.14;
         run_picard.sh CreateSequenceDictionary REFERENCE={input.FASTA} 
OUTPUT={output.DICT} ;
         """

rule sort_sam_picard:
    input:
        BAMREADCOUNT = {rules.bam_read_count.output.TXT},
        BAM = rules.sam_to_bam.output.BAM,
```

```
        DICT = rules.create_seq_dict.output.DICT
    output:
        SortBAM = temporary(root / '{sample}' / 'lofreq' /
'{sample}_picardSorted.bam')
    shell:
         """
         module load picard/2.18.14;
         run_picard.sh SortSam I={input.BAM} O={output.SortBAM}
SORT_ORDER=coordinate
         """


rule index_picardSorted_bam:
    input:
        Dedup_BAM = rules.sort_sam_picard.output.SortBAM
    output:
        index_Dedup_BAM = root / '{sample}' / 'lofreq' /
'{sample}_picardSorted.bam.bai',
    shell:
         """
         module load samtools/1.9;
         samtools index {input.Dedup_BAM}
         """

rule read_groups:
    input:
        Dedup_BAM = rules.sort_sam_picard.output.SortBAM,
        index_Dedup_BAM = rules.index_picardSorted_bam.output.index_Dedup_BAM
    output:
        gp_BAM = temporary(root / '{sample}' / 'lofreq' /  '{sample}_gp.bam'),
    shell:
         """
         module load picard/2.18.14;
         run_picard.sh AddOrReplaceReadGroups I={input.Dedup_BAM} O={output.gp_BAM}
LB=test PL=test PU=test SM=test
         """

rule index_gp_bam:
    input:
        GP_BAM = rules.read_groups.output.gp_BAM
    output:
        index_gp_BAM = root / '{sample}' / 'lofreq' /  '{sample}_gp.bam.bai',
    shell:
         """
         module load samtools/1.9;
         samtools index {input.GP_BAM}
         """

rule realigner_targetcreator:
    input:
        FASTA = rules.consensus_fasta_index.output.INDEX,
        GP_BAM = rules.read_groups.output.gp_BAM,
        index_Dedup_BAM = rules.index_gp_bam.output.index_gp_BAM,
        DICT = rules.create_seq_dict.output.DICT
    output:
        INTERVAL = root / '{sample}' / 'lofreq' /  '{sample}.intervals',
    shell:
         """
         module load gatk/3.7;
         run_gatk.sh -T RealignerTargetCreator -R {input.FASTA} -I {input.GP_BAM} -
o {output.INTERVAL};
```

```
         """

rule indel_realigner:
    input:
        FASTA = rules.consensus_fasta_index.output.INDEX,
        GP_BAM = rules.read_groups.output.gp_BAM,
        INTERVAL = rules.realigner_targetcreator.output.INTERVAL
    output:
        REAL_BAM = temporary(root / '{sample}' / 'lofreq' /  '{sample}_real.bam'),
    shell:
         """
         module load gatk/3.7;
         run_gatk.sh  -T IndelRealigner -maxReads 1000000 -R {input.FASTA} -I
{input.GP_BAM} -targetIntervals {input.INTERVAL} -o {output.REAL_BAM};
         """

rule index_real_bam:
    input:
        REAL_BAM = rules.indel_realigner.output.REAL_BAM
    output:
        index_real_BAM = root / '{sample}' / 'lofreq' /  '{sample}_real.bam.bai',
    shell:
         """
         module load samtools/1.9;
         samtools index {input.REAL_BAM}
         """

rule lofreq_indelqual:
    input:
        FASTA = rules.consensus_fasta_index.output.INDEX,
        REAL_BAM = rules.indel_realigner.output.REAL_BAM,
        INTERVAL = rules.realigner_targetcreator.output.INTERVAL,
        INDEX = rules.index_real_bam.output.index_real_BAM
    output:
        INDELQUAL_BAM = temporary(root / '{sample}' / 'lofreq' /
'{sample}_indelqual.bam'),
    shell:
         """
         module load lofreq_star/2.1.3.1;
         lofreq indelqual --dindel -f {input.FASTA} -o {output.INDELQUAL_BAM}
{input.REAL_BAM}
         """

rule lofreq_indelqual_index:
    input:
        indelqual_BAM = rules.lofreq_indelqual.output.INDELQUAL_BAM
    output:
        index_indelqual_BAM = root / '{sample}' / 'lofreq' /
'{sample}_indelqual.bam.bai',
    shell:
         """
         module load samtools/1.9;
         samtools index {input.indelqual_BAM}
         """

rule lofreq_call:
    input:
        FASTA = rules.consensus_fasta_index.output.INDEX,
        INDELQUAL_BAM = rules.lofreq_indelqual.output.INDELQUAL_BAM,
        index_INDELQUAL_BAM =
rules.lofreq_indelqual_index.output.index_indelqual_BAM
```

```
    output:
        LOFREQ_VCF = root / '{sample}' / 'lofreq' /  '{sample}_unfiltered.vcf',
    shell:
          """
          module load lofreq_star/2.1.3.1;
          lofreq call --call-indels --no-default-filter -f {input.FASTA} -o
{output.LOFREQ_VCF} {input.INDELQUAL_BAM}
          """

rule lofreq_filter:
    input:
        LOFREQ_VCF = rules.lofreq_call.output.LOFREQ_VCF
    output:
        LOFREQ_VCF_2 = root / '{sample}' / 'lofreq' /  '{sample}_filtered.vcf',
    shell:
          """
          module load lofreq_star/2.1.3.1;
          lofreq filter -i {input.LOFREQ_VCF} -o {output.LOFREQ_VCF_2} --sb-thresh
2147483647
          """

rule add_info_to_vcf:
    input:
        REAL_BAM = rules.indel_realigner.output.REAL_BAM,
        LOFREQ_VCF_2 = rules.lofreq_filter.output.LOFREQ_VCF_2
    output:
        POS_Q_VCF_2 = root / '{sample}' / 'lofreq' /  '{sample}_pos_q.vcf'
    params:
        script = os.path.join(snake_dir, "../python/scripts/add_info_to_vcf.py")
    shell:
          """
          python {params.script} {input.REAL_BAM} {input.LOFREQ_VCF_2}
{output.POS_Q_VCF_2}
          """

rule vcf_to_csv:
    input:
        POS_Q_VCF_2 = rules.add_info_to_vcf.output.POS_Q_VCF_2
    output:
        CSV_nF_2 = root / '{sample}' / 'lofreq' /  '{sample}_nofilter.csv'
    params:
        script = os.path.join(snake_dir, "../python/scripts/vcf_to_csv_lofreq.py"),
        base = lambda wildcards: wildcards.sample
    shell:
          """
          python {params.script} -I {input.POS_Q_VCF_2} -O {output.CSV_nF_2} -S
{params.base} -F none
          """
```

### 1 add_info_to_vcf.py

```
# modified from lauringlab (cf
https://github.com/lauringlab/Benchmarking_paper/blob/master/scripts/mapq_vcf.py)
#!/usr/bin/python

import numpy as np
import pysam
import os
import vcf
from vcf.parser import _Info as VcfInfo
import sys


def filter(bam_file=None, in_vcf_file=None, out_vcf_file=None):
    """
    Doesn't filter, per say.
    Adds average phred scores, read positions and MapQ.
    :param bam_file:
    :param in_vcf_file:
    :param out_vcf_file:
    :return:
    """

    # Not useful, it seems.
    # input = in_vcf_file
    # if "/" in input:
    #     input = input.split("/")[-1]
    #
    # input=input.split(".")[0]

    in_var = vcf.Reader(open(in_vcf_file, 'r'))
    ## update infos ##
    in_var.infos['MapQ']=VcfInfo(id='MapQ',num=1,type='Float',desc="The average
MapQ of the reads containing the called variant", source=None, version=None)
    in_var.infos['Read_pos']=VcfInfo(id='Read_pos',num=1,type='Float',desc="The
average read cycle that called the given variant", source=None, version=None)
    in_var.infos['Phred']=VcfInfo(id='Phred',num=1,type='Float',desc="The average
Phred score of the called variant", source=None, version=None)

    variants=list(in_var)
    if len(variants) != 0:
        with pysam.AlignmentFile(bam_file, "rb") as bamfile:

            for record in variants:
                # TD for debugging
                print(record)
                print(record.POS)
                if record.POS == "1026":
                    pass
                mapq=[]  # This will hold a list of the mapping qualities that map
to the variant
                phred=[]  # This will hold a list of the phred that map to the
variant
                Read_pos = []  # This will hold a list of position relative to the
read
                chr=record.CHROM
                pos=int(record.POS)
                py_pos=pos-1
```

```
                var=record.ALT[0]
                # record.ID=input
                #stepper="nofilter", min_base_quality=0,  doesn't change anything
on 1 dataset tested
                for pileupcolumn in
bamfile.pileup(chr,py_pos,py_pos+1,truncate=True,stepper="all", max_depth=1E6):
                    if pileupcolumn.pos==py_pos:
                        for pileupread in pileupcolumn.pileups:
                            if not pileupread.is_del and not pileupread.is_refskip:

called_base=pileupread.alignment.query_sequence[pileupread.query_position]

called_phred=pileupread.alignment.query_qualities[pileupread.query_position]
                                if called_phred>0 and called_base==var: # change
this if you change the phred cut off in deepSNV

mapq.append(pileupread.alignment.mapping_quality)
                                    phred.append(called_phred)
                                    Read_pos.append(pileupread.query_position)
                mean_map=np.mean(mapq)
                mean_phred=np.mean(phred)
                mean_Read_pos=np.mean(Read_pos)

                if mean_map==[]:
                    print( "OOPS didn't find the variant looks like you didn't fix
the bug")
                    sys.exit(1)

                record.add_info('MapQ',mean_map)
                record.add_info('Read_pos',mean_Read_pos)
                record.add_info('Phred',mean_phred)

        print ("done updating")
        iter(variants)
        vcf_writer = vcf.Writer(open(out_vcf_file, 'w'), in_var)
        for record in variants:
            vcf_writer.write_record(record)
        vcf_writer.close()

    else:
        print("No var provided")

def main():
    args = sys.argv[:]
    if len(args) != 4:
        sys.exit("Not all required arguments were present. Exiting.")

    bam_file = args[1]
    in_vcf_file = args[2]
    out_vcf_file = args[3]

    if not os.path.isfile(out_vcf_file):
        if not os.path.isdir(os.path.split(out_vcf_file)[0]):
            os.mkdir(os.path.split(out_vcf_file)[0])
        # print(sample)
        # print(in_vcf_file)
        filter(bam_file, in_vcf_file, out_vcf_file)

if __name__ == main():
    main()
```

#### 2 vcf_to_csv_lofreq.py

```
import csv
import os
import vcf
import sys
import traceback
import argparse


def parse_cla():

    ap = argparse.ArgumentParser()
    ap.add_argument('-O', '--output_csv_path', dest='out_csv_file',
metavar='out_csv_file', required=True, help="Absolute path to output csv file.")
    ap.add_argument('-I', '--in_vcf_path', dest='in_vcf_path',
metavar='in_vcf_path', required=True, help="Absolute path to input vcf file")
    ap.add_argument('-S', '--sample_name', dest='sample_name',
metavar='sample_name', required=True, help="Sample name to be added to each variant
line. Default sampleX", default="sampleX")
    ap.add_argument('-F', '--filter', dest='filter', metavar="filter",
choices=["none", "position", "AF"], help="Filtering to perform; either none (all
variants added to csv), position (only variants within specified average reads
positions are kept), or AF (NOT IMPLEMENTED - filters out based on AF of variant)",
required = True)
    ap.add_argument('-mirp', '--min_read_pos', dest='min_read_pos',
metavar='min_read_pos', required=False, help="Minimal average read position for
filtering on read positions. Required if using --filter position . Default 62.",
default=62)
    ap.add_argument('-marp', '--max_read_pos', dest='max_read_pos',
metavar='max_read_pos', required=False,
                    help="Max average read position for filtering on read
positions. Required if using --filter position . Default 188.",
                    default=188)
    return ap.parse_args()


def validate_args(args=None):
    """
    Check that all provided arguments are correct and coherent, expected files
exist, ....
    :param args:
    :return:
    """
    validated = True

    if args is None:
        print("No args.")
        validated = False
    else:
        if not os.path.isfile(args.in_vcf_path):
            print(f"No vcf file for {args.in_vcf_path}.")
            validated = False

        if args.filter == "AF":
            print(f"filter_high_fq not implemented yet")
            validated = False

    if not validated:
        print(f"Exiting.")
```

```
        sys.exit(1)


def print_pre_run_info(args=None):
    """
    Print warnings and info before running based on args.
    :return:
    """
    print(f"Running with min_read_pos={args.min_read_pos} and max_read_pos =
{args.max_read_pos}.")


def filter_vcf_file(min_read_pos=None, max_read_pos=None, in_vcf_file_path=None,
filter=None):
    """
    Filter the records in vcf file and output the kept records as a list.
    :return: list
    """
    results = []

    with open(in_vcf_file_path, 'r') as in_vcf_file_stream:
        vcf_reader = vcf.Reader(in_vcf_file_stream)
        for record in vcf_reader:
            output_record = [record.CHROM, record.POS, record.REF, record.ALT,
record.INFO["AF"], record.INFO["DP"], record.INFO["SB"]]
            add_record_to_output = False
            try:

                if filter == "none":
                    add_record_to_output = True

                # filter-out extreme average read positions
                elif filter == "position":
                    if min_read_pos < record.INFO['Read_pos'] < max_read_pos:
                        add_record_to_output = True

                # filter on AF values
                elif filter == "AF":
                    pass

                if add_record_to_output:
                    results.append(output_record)

            except:
                print("Error with the following variant:")
                print(output_record)
                print(traceback.format_exc())

    return results

def write_csv(output_csv_path=None, results=None, sample_name=None):
    """
    Write csv file with output records.
    :return:
    """

    out_header = ["Sample", "CHROM", "POS", "REF", "ALT", "VF", "DP", "SB"]

    with open(output_csv_path, "w") as out_csv_stream:
        csv_writer = csv.writer(out_csv_stream)
        csv_writer.writerow(out_header)
```

```python
        for record in results:
            line = [sample_name]
            for elem in record:
                line.append(elem)
            csv_writer.writerow(line)

def main():

    args = parse_cla()

    validate_args(args=args)
    print_pre_run_info(args=args)

    in_vcf_file_path = args.in_vcf_path
    output_csv_path = args.out_csv_file
    sample_name = args.sample_name

    filter = args.filter
    min_read_pos = args.min_read_pos
    max_read_pos = args.max_read_pos

    results = filter_vcf_file(min_read_pos=min_read_pos, max_read_pos=max_read_pos,
in_vcf_file_path=in_vcf_file_path, filter=filter)

    write_csv(output_csv_path=output_csv_path, results=results,
sample_name=sample_name)


main()
```

**Supplementary Table S1: Coverages of the positions characteristic to B.1.1.7 mutations per sample after normalization.** The cells are coloured according to their group, either with a coverage of approximately 5000X (yellow) or approximately 10,000X (green)

| | Mutant | | | | | | | | | | Wild-type | | | | | | | | | |
|---|---|---|---|---|---|---|---|---|---|---|---|---|---|---|---|---|---|---|---|---|
| | ERR5059072 | ERR5059238 | ERR5059260 | ERR5059092 | ERR5059204 | ERR5059123 | ERR5059282 | ERR5058968 | ERR5059226 | ERR5059033 | ERR5059114 | ERR5059253 | ERR5059286 | ERR5059283 | ERR5039162 | ERR5059083 | ERR5059133 | ERR5059257 | ERR5059154 | ERR5040499 |
| **C913T** | 4917 | 4881 | 4845 | 4780 | 4856 | 4837 | 4813 | 4921 | 4850 | 4745 | 4835 | 4862 | 4839 | 4864 | 4929 | 4926 | 4910 | 4828 | 4880 | 4884 |
| **C3267T** | 4849 | 4779 | 4839 | 4814 | 4853 | 4919 | 4864 | 4883 | 4816 | 4870 | 4886 | 4891 | 4786 | 4909 | 4781 | 4896 | 4859 | 4863 | 4839 | 4793 |
| **C5388A** | 4790 | 4809 | 4768 | 4767 | 4886 | 4869 | 4862 | 4803 | 4767 | 4727 | 4867 | 4784 | 4798 | 4902 | 4821 | 4862 | 4861 | 4716 | 4811 | 3580 |
| **C5986T** | 4783 | 4863 | 4878 | 4788 | 4921 | 4940 | 4883 | 4778 | 4872 | 4769 | 4841 | 4904 | 4885 | 4869 | 4907 | 4905 | 4908 | 4847 | 4912 | 4807 |
| **T6954C** | 4793 | 4868 | 4846 | 4746 | 4850 | 4800 | 4839 | 4823 | 4874 | 4724 | 4821 | 4925 | 4849 | 4828 | 4912 | 4856 | 4835 | 4844 | 4872 | 4563 |
| **11288-11296 deletion** | 5015 | 4980 | 4962 | 4904 | 5007 | 4972 | 5034 | 4879 | 4965 | 4944 | 5009 | 4936 | 4992 | 5001 | 5008 | 5040 | 4962 | 4988 | 4973 | 4858 |
| **C14676T** | 4813 | 4825 | 4764 | 4715 | 4834 | 4794 | 4734 | 4861 | 4828 | 4633 | 4780 | 4836 | 4821 | 4763 | 4804 | 4848 | 4870 | 4835 | 4828 | 4731 |
| **C15279T** | 4787 | 4770 | 4777 | 4722 | 4807 | 4831 | 4798 | 4815 | 4798 | 4722 | 4877 | 4789 | 4790 | 4765 | 4855 | 4800 | 4795 | 4797 | 4807 | 4796 |
| **T16176C** | 9712 | 9883 | 9900 | 9489 | 9734 | 9796 | 9783 | 9738 | 9871 | 9627 | 8998 | 9892 | 9855 | 9855 | 9695 | 9817 | 9722 | 9806 | 9898 | 9543 |
| **21765-21770 deletion** | 4846 | 4816 | 4894 | 4729 | 4835 | 4833 | 4795 | 4797 | 4845 | 4688 | 4882 | 4906 | 4803 | 4794 | 4799 | 4841 | 4813 | 4769 | 4882 | 4718 |
| **21991-21993 deletion** | 9718 | 9778 | 9759 | 9552 | 9728 | 9753 | 9766 | 9748 | 9703 | 9494 | 9823 | 9827 | 9774 | 9827 | 9796 | 9723 | 9773 | 9741 | 9850 | 9636 |
| **A23063T** | 4459 | 4912 | 5049 | 4862 | 4839 | 4852 | 4899 | 4886 | 4896 | 4571 | 4916 | 4915 | 4867 | 4928 | 4872 | 4378 | 4458 | 4923 | 4883 | 4759 |
| **C23271A** | 4730 | 4835 | 4713 | 4636 | 4735 | 4823 | 4859 | 4782 | 4772 | 4694 | 4786 | 4830 | 4811 | 4889 | 4776 | 4825 | 4840 | 4796 | 4755 | 4644 |
| **C23604A** | 4807 | 4869 | 4878 | 4766 | 4913 | 4824 | 4822 | 4826 | 4900 | 4710 | 4843 | 4803 | 4860 | 4842 | 4768 | 4868 | 4813 | 4796 | 4932 | 4761 |
| **C23709T** | 4882 | 4904 | 4907 | 4901 | 4952 | 4846 | 4853 | 4852 | 4929 | 4878 | 4868 | 4840 | 4885 | 4869 | 4799 | 4896 | 4846 | 4839 | 4961 | 4781 |
| **T24506G** | 4897 | 4900 | 4969 | 4925 | 4970 | 4882 | 4847 | 4847 | 4976 | 4932 | 4868 | 4827 | 4869 | 4807 | 4830 | 4895 | 4975 | 4949 | 4982 | 4814 |
| **G24914C** | 9428 | 9461 | 9370 | 9244 | 9519 | 9324 | 9458 | 9272 | 9403 | 9063 | 9525 | 9566 | 9460 | 9527 | 9536 | 9545 | 9387 | 9502 | 9459 | 9223 |
| **G26801C** | 4824 | 4829 | 4834 | 4732 | 4805 | 4837 | 4903 | 4854 | 4902 | 4669 | 4803 | 4818 | 4830 | 4870 | 4821 | 4873 | 4870 | 4860 | 4843 | 4770 |
| **C27972T** | 4900 | 4776 | 4851 | 4842 | 4833 | 4868 | 4780 | 4808 | 4877 | 4730 | 9325 | 9507 | 9097 | 9310 | 9304 | 9253 | 9292 | 9311 | 9505 | 7143 |
| **G28048T** | 4879 | 4765 | 4862 | 4750 | 4810 | 4888 | 4759 | 4817 | 4888 | 4679 | 4920 | 4969 | 4791 | 4882 | 4868 | 4888 | 4898 | 4876 | 4880 | 3787 |
| **A28111G** | 9626 | 9671 | 9729 | 9500 | 9655 | 9744 | 9677 | 9653 | 9720 | 9499 | 9719 | 9854 | 9661 | 9767 | 9687 | 9818 | 9852 | 9817 | 9705 | 8570 |
| **G28280C** | 11561 | 10638 | 10424 | 10081 | 10946 | 10522 | 10627 | 10569 | 10227 | 10228 | 10076 | 10285 | 10275 | 9736 | 10389 | 10492 | 10464 | 10299 | 10080 | 10033 |
| **A28281T** | 11563 | 10638 | 10424 | 10080 | 10946 | 10521 | 10626 | 10568 | 10225 | 10230 | 10077 | 10286 | 10276 | 9736 | 10389 | 10492 | 10464 | 10300 | 10079 | 10036 |
| **T28282A** | 11572 | 10638 | 10423 | 10080 | 10947 | 10520 | 10625 | 10568 | 10223 | 10233 | 10078 | 10289 | 10276 | 9736 | 10390 | 10493 | 10464 | 10300 | 10080 | 10037 |
| **C28977T** | 4864 | 4813 | 4845 | 4787 | 4925 | 4914 | 4833 | 4859 | 4835 | 4722 | 4824 | 4953 | 4853 | 4885 | 4782 | 4793 | 4922 | 4780 | 4897 | 4780 |

**Supplementary Table S2: Number of mutations that were considered at which theoretical and median coverage.** If N=7, it indicates that only the group of mutations with an initial coverage of approximately 10,000X was considered. If N=17, only the group of mutations with an initial coverage of approximately 5000X was considered. If N=24, it means that at that coverage all mutations could be considered.

| Number of mutations | Theoretical coverage | Median coverage |
|---|---|---|
| N=7 | 10,000 | 9792 |
| N=7 | 9000 | 8790 |
| N=7 | 8000 | 7801 |
| N=7 | 7000 | 6834 |
| N=7 | 6000 | 5855 |
| N=24 | 5000 | 4855 |
| N=17 | 4500 | 4362 |
| N=24 | 4000 | 3876 |
| N=17 | 3500 | 3386 |
| N=24 | 3000 | 2907 |
| N=17 | 2500 | 2416 |
| N=24 | 2000 | 1939.5 |
| N=24 | 1500 | 1456 |
| N=24 | 1000 | 971 |
| N=17 | 750 | 730 |
| N=24 | 500 | 483 |
| N=17 | 250 | 239 |
| N=7 | 200 | 201 |
| N=17 | 100 | 98 |

**Supplementary Figure S1: Qualitative evaluation of Dataset 1 using the number of false negatives divided by the number of observations per condition until a targeted mutant AF of 100%.** Orange and red dots represent conditions with a FN proportion between 0 and 0.1, and between 0.1 and 1, respectively. The percentage of false negatives is coloured ranging from 0 (dark) to 1 (yellow) in intervals of 0.1 as extrapolated using a contour plot in the R package plotly [61] (actual FN proportions are presented in Supplementary Table S3). Note that a targeted AF of 0% corresponds to the wild-type and is therefore not presented. Both the x- and y-axis follow a logarithmic scale.

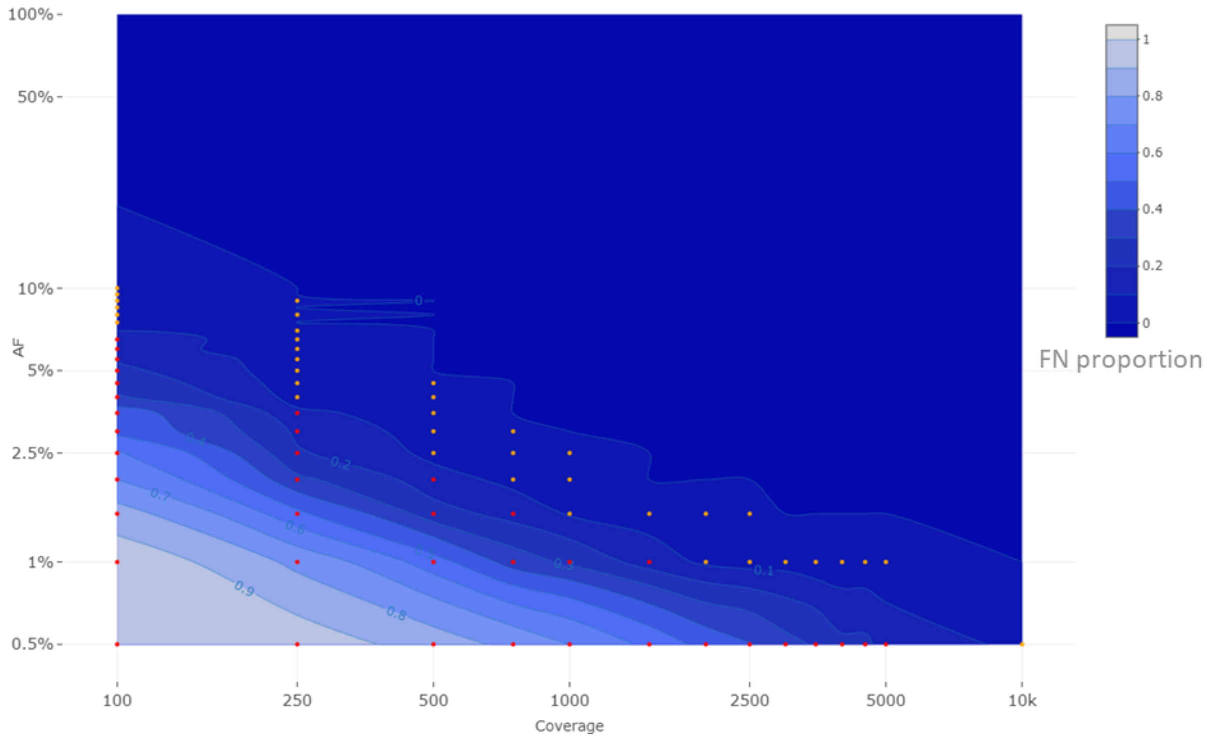

**Supplementary Table S3: Qualitative evaluation of Dataset 1 using the number of false negatives divided by the number of observations per condition until a targeted mutant AF of 100%.** The percentage of false negatives is coloured ranging from 0 (dark) to 1 (light) according to the gradient depicted in

Supplementary Figure **S1**. Actual FN proportions are presented in Table 3. Note that a targeted AF of 0% corresponds to the wild-type and it is therefore not presented.

| **Coverage →**<br>**AF ↓** | 100 | 250 | 500 | 750 | 1000 | 1500 | 2000 | 2500 | 3000 | 3500 | 4000 | 4500 | 5000 | 10,000 |
|---|---|---|---|---|---|---|---|---|---|---|---|---|---|---|
| 100.0% | 0% | 0% | 0% | 0% | 0% | 0% | 0% | 0% | 0% | 0% | 0% | 0% | 0% | 0% |
| 50.0% | 0% | 0% | 0% | 0% | 0% | 0% | 0% | 0% | 0% | 0% | 0% | 0% | 0% | 0% |
| 40.0% | 0% | 0% | 0% | 0% | 0% | 0% | 0% | 0% | 0% | 0% | 0% | 0% | 0% | 0% |
| 30.0% | 0% | 0% | 0% | 0% | 0% | 0% | 0% | 0% | 0% | 0% | 0% | 0% | 0% | 0% |
| 20.0% | 0% | 0% | 0% | 0% | 0% | 0% | 0% | 0% | 0% | 0% | 0% | 0% | 0% | 0% |
| 10.0% | 5% | 0% | 0% | 0% | 0% | 0% | 0% | 0% | 0% | 0% | 0% | 0% | 0% | 0% |
| 9.5% | 4% | 0% | 0% | 0% | 0% | 0% | 0% | 0% | 0% | 0% | 0% | 0% | 0% | 0% |
| 9.0% | 7% | 1% | 0% | 0% | 0% | 0% | 0% | 0% | 0% | 0% | 0% | 0% | 0% | 0% |
| 8.5% | 4% | 0% | 0% | 0% | 0% | 0% | 0% | 0% | 0% | 0% | 0% | 0% | 0% | 0% |
| 8.0% | 9% | 2% | 0% | 0% | 0% | 0% | 0% | 0% | 0% | 0% | 0% | 0% | 0% | 0% |
| 7.5% | 8% | 0% | 0% | 0% | 0% | 0% | 0% | 0% | 0% | 0% | 0% | 0% | 0% | 0% |
| 7.0% | 10% | 1% | 0% | 0% | 0% | 0% | 0% | 0% | 0% | 0% | 0% | 0% | 0% | 0% |
| 6.5% | 15% | 2% | 0% | 0% | 0% | 0% | 0% | 0% | 0% | 0% | 0% | 0% | 0% | 0% |
| 6.0% | 15% | 1% | 0% | 0% | 0% | 0% | 0% | 0% | 0% | 0% | 0% | 0% | 0% | 0% |
| 5.5% | 19% | 3% | 0% | 0% | 0% | 0% | 0% | 0% | 0% | 0% | 0% | 0% | 0% | 0% |
| 5.0% | 22% | 3% | 0% | 0% | 0% | 0% | 0% | 0% | 0% | 0% | 0% | 0% | 0% | 0% |
| 4.5% | 26% | 4% | 2% | 0% | 0% | 0% | 0% | 0% | 0% | 0% | 0% | 0% | 0% | 0% |
| 4.0% | 31% | 6% | 1% | 0% | 0% | 0% | 0% | 0% | 0% | 0% | 0% | 0% | 0% | 0% |
| 3.5% | 45% | 12% | 4% | 0% | 0% | 0% | 0% | 0% | 0% | 0% | 0% | 0% | 0% | 0% |
| 3.0% | 47% | 18% | 4% | 1% | 0% | 0% | 0% | 0% | 0% | 0% | 0% | 0% | 0% | 0% |
| 2.5% | 62% | 21% | 7% | 2% | 2% | 0% | 0% | 0% | 0% | 0% | 0% | 0% | 0% | 0% |
| 2.0% | 70% | 32% | 14% | 7% | 3% | 0% | 0% | 0% | 0% | 0% | 0% | 0% | 0% | 0% |
| 1.5% | 84% | 52% | 24% | 16% | 9% | 5% | 1% | 2% | 0% | 0% | 0% | 0% | 0% | 0% |
| 1.0% | 96% | 77% | 54% | 35% | 28% | 15% | 8% | 6% | 6% | 3% | 2% | 2% | 2% | 0% |
| 0.5% | 98% | 95% | 85% | 77% | 70% | 57% | 46% | 41% | 33% | 29% | 22% | 22% | 16% | 7% |
| 0.0% | 0% | 0% | 0% | 0% | 0% | 0% | 0% | 0% | 0% | 0% | 0% | 0% | 0% | 0% |

**Supplementary Figure S2: Qualitative evaluation of Dataset 2 using the number of false negatives divided by the number of observations per condition until a targeted mutant AF of 100%.** Orange and red dots represent conditions with a FN proportion between 0 and 0.1, and between 0.1 and 1, respectively. The percentage of false negatives is coloured ranging from 0 (dark) to 1 (light) in intervals of 0.1 as extrapolated using a contour plot in the R package plotly [61] (actual FN proportions are presented in Supplementary Table S4). Note that a targeted AF of 0% represents to the wild-type and is therefore not presented. Both the x- and y-axis follow a logarithmic scale.

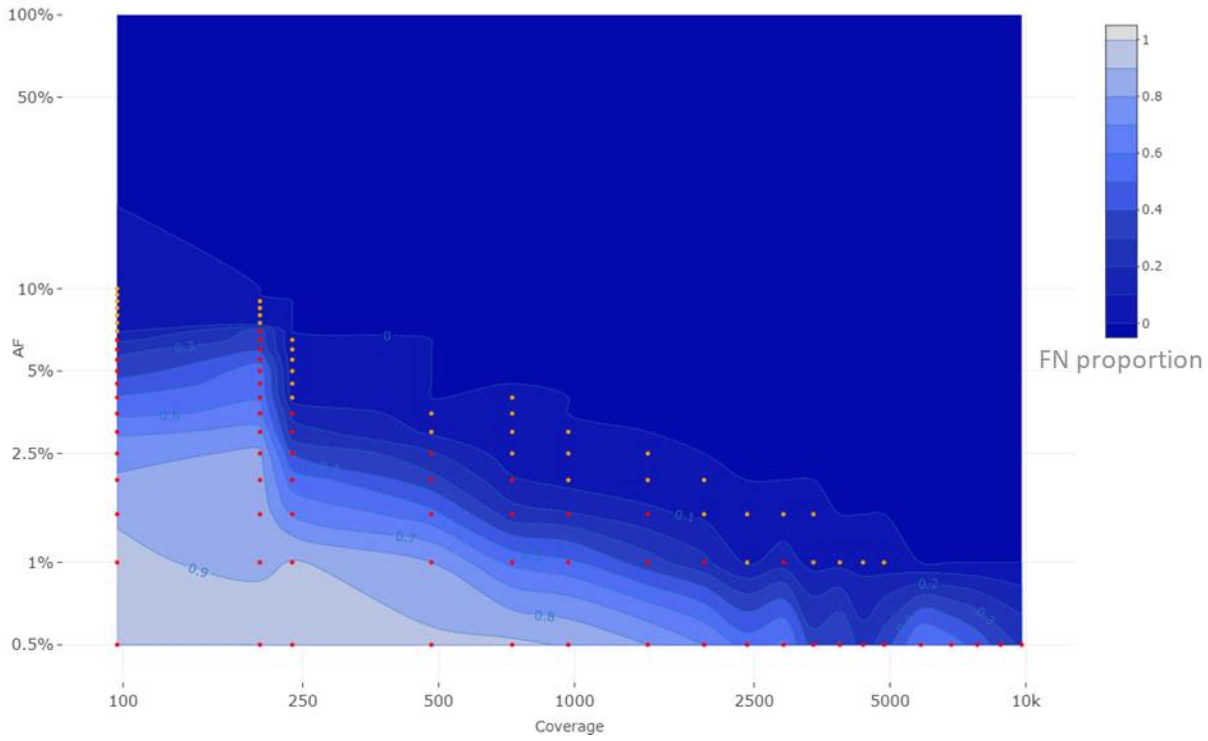

**Supplementary Table S4: Qualitative evaluation of Dataset 2 using the number of false negatives divided by the number of observations per condition until a targeted mutant AF of 100%.** The percentage of false negatives is coloured ranging from 0 (dark) to 1 (light) according to the gradient depicted in Supplementary Figure S2. Actual FN proportions are presented in Table 3. Note that a targeted AF of 0% corresponds to the wild-type and it is therefore not presented.

| Coverage → AF ↓ | 97 | 201 | 237 | 482 | 728 | 969 | 1454 | 1937 | 2413 | 2904 | 3383 | 3872 | 4358 | 4851 | 5855 | 6834 | 7801 | 8790 | 9792 |
|---|---|---|---|---|---|---|---|---|---|---|---|---|---|---|---|---|---|---|---|
| 100.0% | 0% | 0% | 0% | 0% | 0% | 0% | 0% | 0% | 0% | 0% | 0% | 0% | 0% | 0% | 0% | 0% | 0% | 0% | 0% |
| 50.0% | 0% | 0% | 0% | 0% | 0% | 0% | 0% | 0% | 0% | 0% | 0% | 0% | 0% | 0% | 0% | 0% | 0% | 0% | 0% |
| 40.0% | 0% | 0% | 0% | 0% | 0% | 0% | 0% | 0% | 0% | 0% | 0% | 0% | 0% | 0% | 0% | 0% | 0% | 0% | 0% |
| 30.0% | 0% | 0% | 0% | 0% | 0% | 0% | 0% | 0% | 0% | 0% | 0% | 0% | 0% | 0% | 0% | 0% | 0% | 0% | 0% |
| 20.0% | 0% | 0% | 0% | 0% | 0% | 0% | 0% | 0% | 0% | 0% | 0% | 0% | 0% | 0% | 0% | 0% | 0% | 0% | 0% |
| 10.0% | 2% | 0% | 0% | 0% | 0% | 0% | 0% | 0% | 0% | 0% | 0% | 0% | 0% | 0% | 0% | 0% | 0% | 0% | 0% |
| 9.5% | 3% | 0% | 0% | 0% | 0% | 0% | 0% | 0% | 0% | 0% | 0% | 0% | 0% | 0% | 0% | 0% | 0% | 0% | 0% |
| 9.0% | 5% | 1% | 0% | 0% | 0% | 0% | 0% | 0% | 0% | 0% | 0% | 0% | 0% | 0% | 0% | 0% | 0% | 0% | 0% |
| 8.5% | 6% | 1% | 0% | 0% | 0% | 0% | 0% | 0% | 0% | 0% | 0% | 0% | 0% | 0% | 0% | 0% | 0% | 0% | 0% |
| 8.0% | 8% | 1% | 0% | 0% | 0% | 0% | 0% | 0% | 0% | 0% | 0% | 0% | 0% | 0% | 0% | 0% | 0% | 0% | 0% |
| 7.5% | 8% | 1% | 0% | 0% | 0% | 0% | 0% | 0% | 0% | 0% | 0% | 0% | 0% | 0% | 0% | 0% | 0% | 0% | 0% |
| 7.0% | 9% | 34% | 0% | 0% | 0% | 0% | 0% | 0% | 0% | 0% | 0% | 0% | 0% | 0% | 0% | 0% | 0% | 0% | 0% |
| 6.5% | 18% | 35% | 1% | 0% | 0% | 0% | 0% | 0% | 0% | 0% | 0% | 0% | 0% | 0% | 0% | 0% | 0% | 0% | 0% |
| 6.0% | 28% | 38% | 1% | 0% | 0% | 0% | 0% | 0% | 0% | 0% | 0% | 0% | 0% | 0% | 0% | 0% | 0% | 0% | 0% |
| 5.5% | 31% | 47% | 3% | 0% | 0% | 0% | 0% | 0% | 0% | 0% | 0% | 0% | 0% | 0% | 0% | 0% | 0% | 0% | 0% |
| 5.0% | 35% | 56% | 3% | 0% | 0% | 0% | 0% | 0% | 0% | 0% | 0% | 0% | 0% | 0% | 0% | 0% | 0% | 0% | 0% |
| 4.5% | 43% | 57% | 4% | 0% | 0% | 0% | 0% | 0% | 0% | 0% | 0% | 0% | 0% | 0% | 0% | 0% | 0% | 0% | 0% |
| 4.0% | 51% | 59% | 6% | 0% | 1% | 0% | 0% | 0% | 0% | 0% | 0% | 0% | 0% | 0% | 0% | 0% | 0% | 0% | 0% |
| 3.5% | 58% | 63% | 18% | 4% | 1% | 0% | 0% | 0% | 0% | 0% | 0% | 0% | 0% | 0% | 0% | 0% | 0% | 0% | 0% |
| 3.0% | 68% | 73% | 23% | 8% | 2% | 1% | 0% | 0% | 0% | 0% | 0% | 0% | 0% | 0% | 0% | 0% | 0% | 0% | 0% |
| 2.5% | 77% | 82% | 40% | 21% | 4% | 3% | 1% | 0% | 0% | 0% | 0% | 0% | 0% | 0% | 0% | 0% | 0% | 0% | 0% |
| 2.0% | 81% | 84% | 55% | 33% | 11% | 6% | 4% | 1% | 0% | 0% | 0% | 0% | 0% | 0% | 0% | 0% | 0% | 0% | 0% |
| 1.5% | 89% | 86% | 69% | 53% | 24% | 21% | 12% | 8% | 4% | 2% | 1% | 0% | 0% | 0% | 0% | 0% | 0% | 0% | 0% |
| 1.0% | 92% | 86% | 91% | 80% | 57% | 52% | 34% | 22% | 8% | 15% | 6% | 7% | 6% | 4% | 0% | 0% | 0% | 0% | 0% |
| 0.5% | 100% | 98% | 98% | 92% | 92% | 89% | 80% | 70% | 55% | 62% | 34% | 41% | 24% | 35% | 62% | 55% | 46% | 35% | 28% |
| 0.0% | 0% | 0% | 0% | 0% | 0% | 0% | 0% | 0% | 0% | 0% | 0% | 0% | 0% | 0% | 0% | 0% | 0% | 0% | 0% |

**Supplementary File S3: Variation of Dataset 1.** In the plots of A and B the SD on the y-axis is plotted against the AF (Reference value) on the x-axis (A: range 0-50%; B: range 0-10%) for each coverage (colours). Table C includes the actual values for the performance metric that was visualized in Figure 3A. In table D the interquartile ranges for each condition is indicated. In the tables, conditions with a false negative percentage greater than 75% were excluded. Note that a targeted AF of 0% and 100% corresponds to the wild-type and mutant respectively and is therefore not presented.

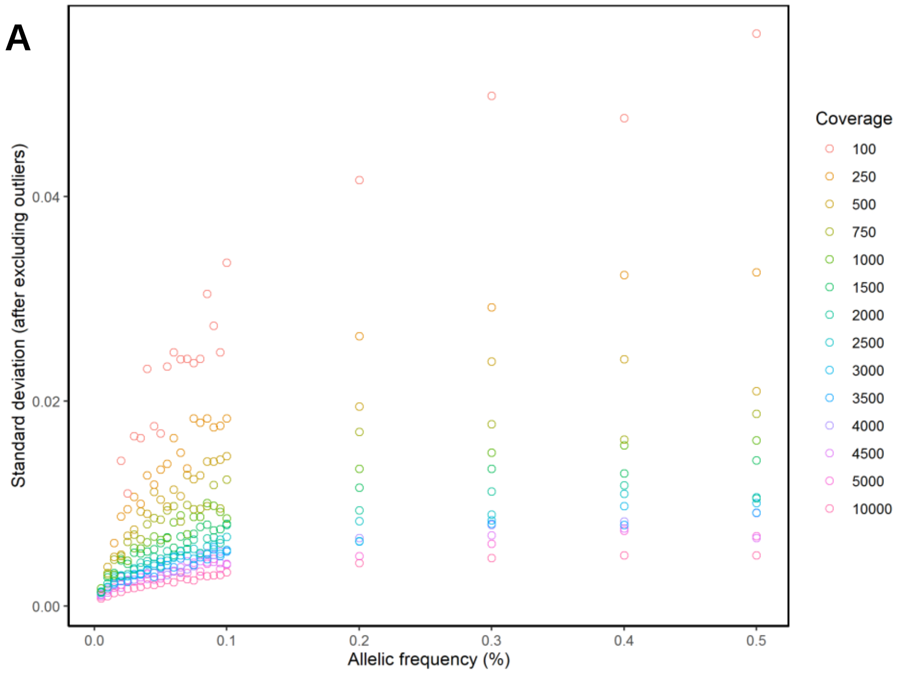


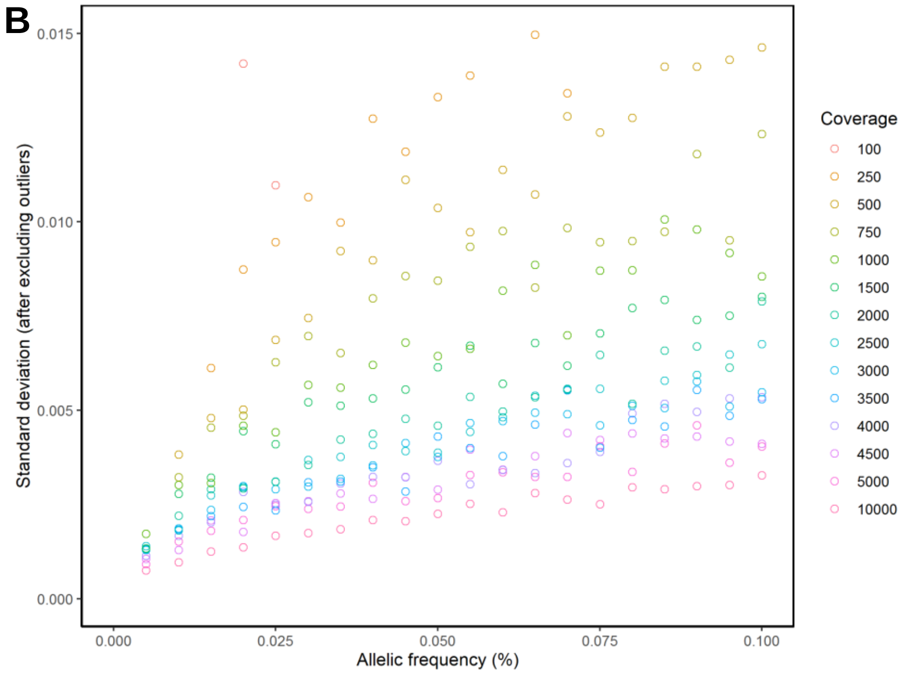

C

| Coverage → AF ↓ | 100 | 250 | 500 | 750 | 1000 | 1500 | 2000 | 2500 | 3000 | 3500 | 4000 | 4500 | 5000 | 10000 |
|---|---|---|---|---|---|---|---|---|---|---|---|---|---|---|
| **50.0%** | 1.00 | 0.34 | 0.14 | 0.11 | 0.08 | 0.06 | 0.04 | 0.04 | 0.03 | 0.03 | 0.03 | 0.01 | 0.01 | 0.01 |
| **40.0%** | 1.00 | 0.46 | 0.26 | 0.12 | 0.11 | 0.07 | 0.06 | 0.05 | 0.04 | 0.03 | 0.03 | 0.03 | 0.02 | 0.01 |
| **30.0%** | 1.00 | 0.34 | 0.23 | 0.13 | 0.09 | 0.07 | 0.05 | 0.03 | 0.03 | 0.03 | 0.03 | 0.02 | 0.01 | 0.01 |
| **20.0%** | 1.00 | 0.40 | 0.22 | 0.17 | 0.10 | 0.08 | 0.05 | 0.04 | 0.02 | 0.02 | 0.03 | 0.03 | 0.01 | 0.01 |
| **10.0%** | 1.00 | 0.30 | 0.19 | 0.14 | 0.06 | 0.06 | 0.06 | 0.04 | 0.03 | 0.02 | 0.03 | 0.01 | 0.01 | 0.01 |
| **9.5%** | 1.00 | 0.50 | 0.33 | 0.15 | 0.14 | 0.09 | 0.06 | 0.07 | 0.04 | 0.04 | 0.05 | 0.03 | 0.02 | 0.01 |
| **9.0%** | 1.00 | 0.41 | 0.27 | 0.19 | 0.13 | 0.07 | 0.06 | 0.05 | 0.04 | 0.04 | 0.03 | 0.02 | 0.03 | 0.01 |
| **8.5%** | 1.00 | 0.36 | 0.21 | 0.10 | 0.11 | 0.07 | 0.05 | 0.04 | 0.03 | 0.02 | 0.03 | 0.02 | 0.02 | 0.01 |
| **8.0%** | 1.00 | 0.55 | 0.28 | 0.15 | 0.13 | 0.10 | 0.05 | 0.05 | 0.05 | 0.04 | 0.04 | 0.03 | 0.02 | 0.02 |
| **7.5%** | 1.00 | 0.60 | 0.27 | 0.16 | 0.13 | 0.09 | 0.07 | 0.06 | 0.04 | 0.03 | 0.03 | 0.03 | 0.03 | 0.01 |
| **7.0%** | 1.00 | 0.31 | 0.28 | 0.17 | 0.08 | 0.07 | 0.05 | 0.05 | 0.04 | 0.05 | 0.02 | 0.03 | 0.02 | 0.01 |
| **6.5%** | 1.00 | 0.39 | 0.20 | 0.12 | 0.14 | 0.08 | 0.05 | 0.05 | 0.04 | 0.04 | 0.02 | 0.02 | 0.02 | 0.01 |
| **6.0%** | 1.00 | 0.44 | 0.21 | 0.16 | 0.11 | 0.05 | 0.04 | 0.04 | 0.04 | 0.02 | 0.02 | 0.02 | 0.02 | 0.01 |
| **5.5%** | 1.00 | 0.35 | 0.17 | 0.16 | 0.08 | 0.08 | 0.05 | 0.04 | 0.04 | 0.03 | 0.02 | 0.03 | 0.02 | 0.01 |
| **5.0%** | 1.00 | 0.63 | 0.38 | 0.25 | 0.15 | 0.13 | 0.07 | 0.05 | 0.05 | 0.07 | 0.05 | 0.03 | 0.03 | 0.02 |
| **4.5%** | 1.00 | 0.46 | 0.40 | 0.24 | 0.15 | 0.10 | 0.07 | 0.05 | 0.06 | 0.03 | 0.03 | 0.03 | 0.02 | 0.01 |
| **4.0%** | 1.00 | 0.30 | 0.15 | 0.12 | 0.07 | 0.05 | 0.04 | 0.03 | 0.02 | 0.02 | 0.02 | 0.01 | 0.02 | 0.01 |
| **3.5%** | 1.00 | 0.37 | 0.32 | 0.16 | 0.12 | 0.10 | 0.07 | 0.05 | 0.04 | 0.04 | 0.03 | 0.03 | 0.02 | 0.01 |
| **3.0%** | 1.00 | 0.41 | 0.20 | 0.18 | 0.12 | 0.10 | 0.05 | 0.05 | 0.03 | 0.03 | 0.02 | 0.02 | 0.02 | 0.01 |
| **2.5%** | 1.00 | 0.74 | 0.39 | 0.33 | 0.16 | 0.14 | 0.08 | 0.08 | 0.07 | 0.05 | 0.05 | 0.05 | 0.05 | 0.02 |
| **2.0%** | 1.00 | 0.38 | 0.13 | 0.12 | 0.10 | 0.10 | 0.04 | 0.04 | 0.04 | 0.03 | 0.04 | 0.02 | 0.02 | 0.01 |
| **1.5%** | | 1.00 | 0.61 | 0.55 | 0.25 | 0.28 | 0.23 | 0.20 | 0.15 | 0.13 | 0.11 | 0.12 | 0.09 | 0.04 |
| **1.0%** | | | 1.00 | 0.71 | 0.63 | 0.53 | 0.33 | 0.23 | 0.24 | 0.23 | 0.19 | 0.11 | 0.16 | 0.06 |
| **0.5%** | | | | | 1.00 | 0.59 | 0.59 | 0.65 | 0.56 | 0.57 | 0.44 | 0.38 | 0.28 | 0.19 |

## D

| Coverage → AF ↓ | 100 | 250 | 500 | 750 | 1000 | 1500 | 2000 | 2500 | 3000 | 3500 | 4000 | 4500 | 5000 | 10,000 |
|---|---|---|---|---|---|---|---|---|---|---|---|---|---|---|
| **50.0%** | 6.26% | 4.10% | 2.44% | 2.06% | 1.93% | 1.79% | 1.28% | 1.39% | 1.28% | 0.97% | 0.91% | 0.88% | 0.70% | 0.62% |
| **40.0%** | 5.86% | 3.45% | 2.37% | 1.94% | 1.98% | 1.55% | 1.33% | 1.18% | 1.17% | 0.87% | 1.00% | 0.91% | 0.79% | 0.55% |
| **30.0%** | 6.06% | 3.40% | 2.83% | 2.02% | 1.89% | 1.36% | 1.39% | 1.21% | 1.07% | 0.97% | 0.84% | 0.88% | 0.86% | 0.55% |
| **20.0%** | 4.59% | 2.74% | 2.26% | 1.99% | 1.47% | 1.34% | 1.05% | 0.90% | 0.89% | 0.95% | 0.73% | 0.74% | 0.67% | 0.52% |
| **10.0%** | 3.49% | 2.47% | 1.65% | 1.33% | 0.98% | 0.85% | 0.83% | 0.86% | 0.70% | 0.59% | 0.60% | 0.50% | 0.55% | 0.36% |
| **9.5%** | 3.03% | 2.23% | 1.81% | 1.44% | 1.00% | 0.71% | 0.80% | 0.67% | 0.74% | 0.67% | 0.56% | 0.48% | 0.42% | 0.37% |
| **9.0%** | 3.16% | 2.11% | 1.65% | 1.27% | 1.14% | 0.91% | 0.78% | 0.73% | 0.67% | 0.62% | 0.56% | 0.55% | 0.47% | 0.34% |
| **8.5%** | 3.50% | 2.09% | 1.63% | 1.25% | 0.90% | 0.92% | 0.69% | 0.65% | 0.61% | 0.54% | 0.62% | 0.48% | 0.45% | 0.31% |
| **8.0%** | 2.95% | 1.81% | 1.53% | 1.24% | 1.03% | 0.87% | 0.68% | 0.56% | 0.52% | 0.47% | 0.52% | 0.54% | 0.48% | 0.32% |
| **7.5%** | 3.12% | 2.02% | 1.55% | 1.14% | 0.96% | 0.84% | 0.74% | 0.67% | 0.60% | 0.50% | 0.59% | 0.52% | 0.51% | 0.31% |
| **7.0%** | 3.15% | 2.26% | 1.49% | 1.16% | 0.92% | 0.80% | 0.71% | 0.53% | 0.61% | 0.62% | 0.50% | 0.48% | 0.51% | 0.29% |
| **6.5%** | 2.38% | 1.64% | 1.42% | 1.25% | 0.90% | 0.78% | 0.59% | 0.62% | 0.53% | 0.53% | 0.41% | 0.47% | 0.40% | 0.33% |
| **6.0%** | 2.87% | 1.87% | 1.34% | 1.17% | 0.90% | 0.73% | 0.71% | 0.55% | 0.50% | 0.51% | 0.40% | 0.44% | 0.41% | 0.26% |
| **5.5%** | 2.47% | 1.60% | 1.06% | 1.12% | 0.84% | 0.69% | 0.59% | 0.46% | 0.48% | 0.47% | 0.48% | 0.44% | 0.39% | 0.28% |
| **5.0%** | 2.07% | 1.55% | 1.32% | 0.97% | 0.67% | 0.70% | 0.52% | 0.55% | 0.47% | 0.51% | 0.44% | 0.39% | 0.30% | 0.27% |
| **4.5%** | 2.41% | 1.58% | 1.15% | 0.90% | 0.87% | 0.64% | 0.58% | 0.48% | 0.46% | 0.38% | 0.37% | 0.35% | 0.30% | 0.25% |
| **4.0%** | 2.41% | 1.42% | 1.18% | 0.92% | 0.68% | 0.58% | 0.53% | 0.47% | 0.48% | 0.42% | 0.36% | 0.38% | 0.34% | 0.25% |
| **3.5%** | 1.81% | 1.24% | 1.04% | 0.75% | 0.73% | 0.59% | 0.55% | 0.41% | 0.38% | 0.37% | 0.40% | 0.29% | 0.32% | 0.23% |
| **3.0%** | 2.02% | 1.28% | 0.83% | 0.86% | 0.76% | 0.67% | 0.49% | 0.42% | 0.38% | 0.38% | 0.29% | 0.30% | 0.31% | 0.20% |
| **2.5%** | 2.08% | 1.37% | 0.78% | 0.74% | 0.59% | 0.47% | 0.37% | 0.37% | 0.29% | 0.34% | 0.33% | 0.28% | 0.30% | 0.21% |
| **2.0%** | 2.08% | 1.13% | 0.70% | 0.56% | 0.52% | 0.41% | 0.33% | 0.37% | 0.33% | 0.29% | 0.31% | 0.27% | 0.22% | 0.20% |
| **1.5%** | 1.65% | 0.86% | 0.57% | 0.48% | 0.37% | 0.33% | 0.36% | 0.28% | 0.28% | 0.26% | 0.25% | 0.22% | 0.22% | 0.14% |
| **1.0%** | 1.82% | 0.92% | 0.54% | 0.39% | 0.30% | 0.25% | 0.31% | 0.22% | 0.21% | 0.22% | 0.20% | 0.17% | 0.18% | 0.11% |
| **0.5%** | 1.16% | 0.99% | 0.60% | 0.41% | 0.33% | 0.24% | 0.17% | 0.16% | 0.14% | 0.13% | 0.14% | 0.12% | 0.10% | 0.08% |

**Supplementary File S4: Variation of Dataset 2.** In the plots of A and B the SD on the y-axis is plotted against the AF on the x-axis (A: range 0-50%; B: range 0-10%) for each coverage (colours). Table C includes the actual values for the performance metric that was visualized in Figure 3B. In table D the interquartile ranges for each condition is indicated, while excluding false negative results. In the tables, conditions with a false negative percentage greater than 75% were excluded. Note that a targeted AF of 0% and 100% corresponds to the wild-type and mutant respectively and is therefore not presented.

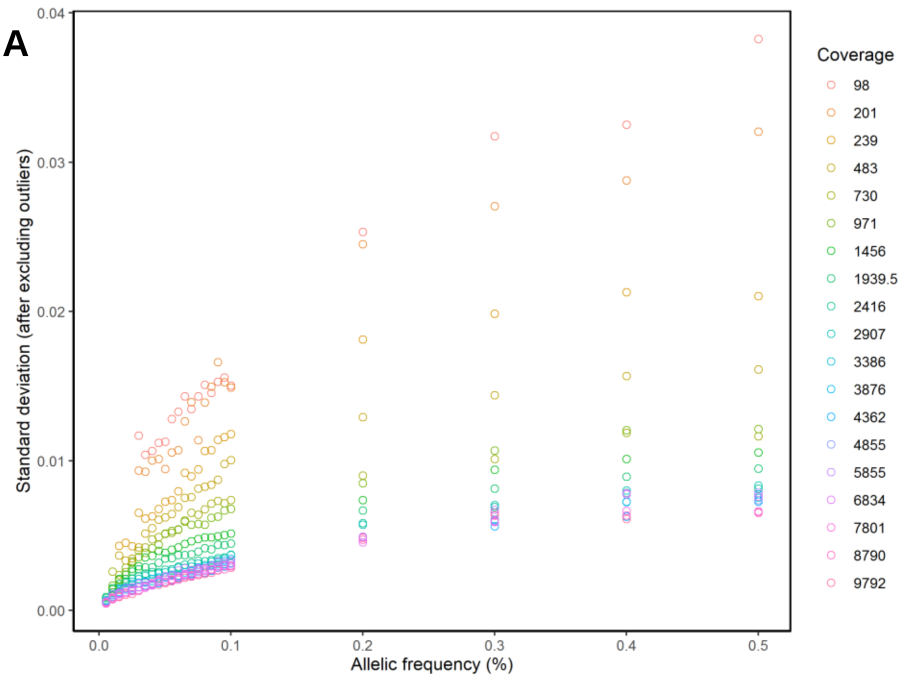


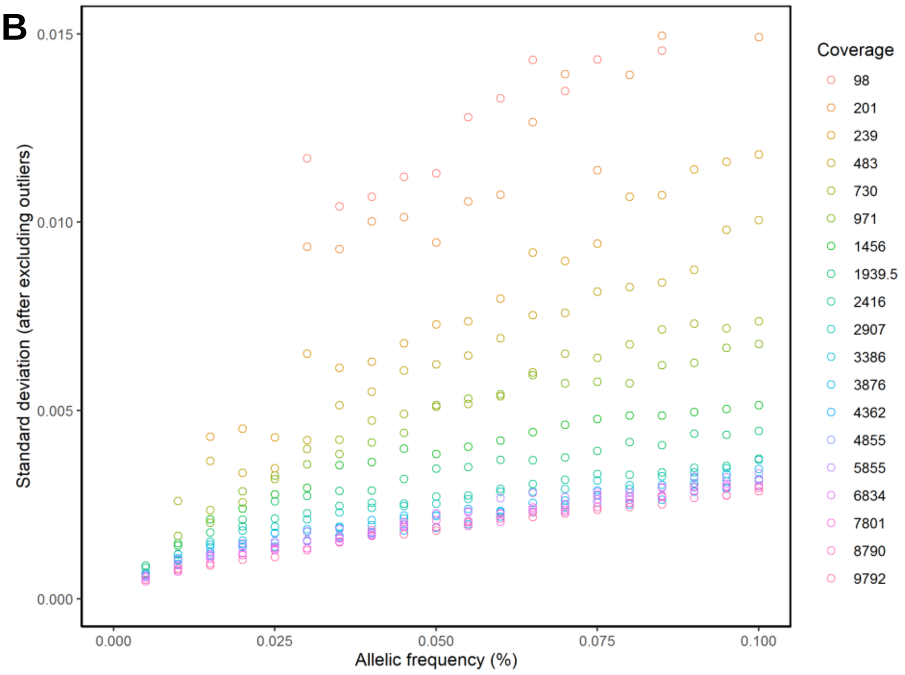

C

| Coverage → AF ↓ | 97 | 201 | 237 | 482 | 728 | 969 | 1454 | 1937 | 2413 | 2904 | 3383 | 3872 | 4358 | 4851 | 5855 | 6834 | 7801 | 8790 | 9792 |
|---|---|---|---|---|---|---|---|---|---|---|---|---|---|---|---|---|---|---|---|
| **50.0%** | 1.00 | 0.70 | 0.30 | 0.18 | 0.09 | 0.10 | 0.08 | 0.06 | 0.05 | 0.05 | 0.04 | 0.04 | 0.04 | 0.04 | 0.04 | 0.04 | 0.03 | 0.03 | 0.03 |
| **40.0%** | 1.00 | 0.78 | 0.43 | 0.23 | 0.13 | 0.14 | 0.10 | 0.08 | 0.06 | 0.06 | 0.05 | 0.05 | 0.04 | 0.04 | 0.06 | 0.06 | 0.04 | 0.04 | 0.04 |
| **30.0%** | 1.00 | 0.73 | 0.39 | 0.21 | 0.10 | 0.11 | 0.09 | 0.07 | 0.05 | 0.05 | 0.04 | 0.04 | 0.03 | 0.03 | 0.05 | 0.04 | 0.03 | 0.04 | 0.04 |
| **20.0%** | 1.00 | 0.93 | 0.51 | 0.26 | 0.13 | 0.11 | 0.08 | 0.07 | 0.05 | 0.05 | 0.03 | 0.04 | 0.03 | 0.03 | 0.04 | 0.03 | 0.04 | 0.03 | 0.04 |
| **10.0%** | 1.00 | 0.98 | 0.62 | 0.45 | 0.24 | 0.20 | 0.12 | 0.09 | 0.06 | 0.06 | 0.04 | 0.05 | 0.04 | 0.04 | 0.05 | 0.04 | 0.04 | 0.04 | 0.04 |
| **9.5%** | 1.00 | 0.96 | 0.56 | 0.40 | 0.21 | 0.18 | 0.10 | 0.08 | 0.05 | 0.05 | 0.04 | 0.05 | 0.04 | 0.04 | 0.04 | 0.04 | 0.04 | 0.03 | 0.03 |
| **9.0%** | 0.85 | 1.00 | 0.47 | 0.28 | 0.19 | 0.14 | 0.09 | 0.07 | 0.04 | 0.04 | 0.03 | 0.04 | 0.03 | 0.03 | 0.04 | 0.03 | 0.03 | 0.03 | 0.03 |
| **8.5%** | 0.95 | 1.00 | 0.51 | 0.32 | 0.23 | 0.17 | 0.11 | 0.07 | 0.05 | 0.05 | 0.04 | 0.04 | 0.03 | 0.03 | 0.04 | 0.04 | 0.03 | 0.03 | 0.03 |
| **8.0%** | 1.00 | 0.85 | 0.50 | 0.30 | 0.20 | 0.14 | 0.10 | 0.08 | 0.05 | 0.04 | 0.03 | 0.04 | 0.03 | 0.03 | 0.03 | 0.03 | 0.03 | 0.03 | 0.03 |
| **7.5%** | 1.00 | 0.63 | 0.43 | 0.32 | 0.20 | 0.16 | 0.11 | 0.08 | 0.05 | 0.05 | 0.04 | 0.04 | 0.03 | 0.03 | 0.04 | 0.04 | 0.03 | 0.03 | 0.03 |
| **7.0%** | 0.94 | 1.00 | 0.41 | 0.30 | 0.22 | 0.17 | 0.11 | 0.07 | 0.05 | 0.04 | 0.03 | 0.04 | 0.03 | 0.03 | 0.04 | 0.03 | 0.03 | 0.03 | 0.03 |
| **6.5%** | 1.00 | 0.78 | 0.41 | 0.28 | 0.18 | 0.17 | 0.10 | 0.07 | 0.05 | 0.04 | 0.03 | 0.03 | 0.03 | 0.03 | 0.04 | 0.03 | 0.03 | 0.03 | 0.02 |
| **6.0%** | 1.00 | 0.65 | 0.36 | 0.27 | 0.16 | 0.17 | 0.10 | 0.08 | 0.05 | 0.05 | 0.03 | 0.03 | 0.03 | 0.03 | 0.04 | 0.03 | 0.02 | 0.03 | 0.03 |
| **5.5%** | 1.00 | 0.68 | 0.33 | 0.25 | 0.16 | 0.17 | 0.10 | 0.07 | 0.05 | 0.04 | 0.03 | 0.03 | 0.02 | 0.03 | 0.03 | 0.03 | 0.03 | 0.02 | 0.03 |
| **5.0%** | 1.00 | 0.70 | 0.42 | 0.30 | 0.20 | 0.21 | 0.12 | 0.09 | 0.06 | 0.05 | 0.04 | 0.04 | 0.03 | 0.03 | 0.04 | 0.04 | 0.03 | 0.03 | 0.03 |
| **4.5%** | 1.00 | 0.82 | 0.37 | 0.29 | 0.19 | 0.15 | 0.13 | 0.08 | 0.05 | 0.05 | 0.04 | 0.04 | 0.03 | 0.03 | 0.03 | 0.03 | 0.03 | 0.03 | 0.02 |
| **4.0%** | 1.00 | 0.88 | 0.35 | 0.27 | 0.20 | 0.15 | 0.12 | 0.07 | 0.06 | 0.05 | 0.03 | 0.04 | 0.03 | 0.03 | 0.03 | 0.03 | 0.02 | 0.03 | 0.02 |
| **3.5%** | 1.00 | 0.80 | 0.35 | 0.24 | 0.16 | 0.14 | 0.12 | 0.08 | 0.06 | 0.05 | 0.03 | 0.03 | 0.03 | 0.02 | 0.03 | 0.02 | 0.02 | 0.02 | 0.02 |
| **3.0%** | 1.00 | 0.64 | 0.31 | 0.13 | 0.12 | 0.09 | 0.06 | 0.05 | 0.04 | 0.03 | 0.02 | 0.02 | 0.02 | 0.02 | 0.02 | 0.02 | 0.01 | 0.01 | 0.01 |
| **2.5%** | | | 1.00 | 0.65 | 0.55 | 0.58 | 0.42 | 0.36 | 0.25 | 0.20 | 0.17 | 0.16 | 0.11 | 0.12 | 0.13 | 0.09 | 0.09 | 0.10 | 0.07 |
| **2.0%** | | | 1.00 | 0.55 | 0.32 | 0.40 | 0.28 | 0.22 | 0.18 | 0.16 | 0.12 | 0.10 | 0.09 | 0.09 | 0.11 | 0.07 | 0.07 | 0.07 | 0.05 |
| **1.5%** | | | 1.00 | 0.73 | 0.30 | 0.22 | 0.24 | 0.17 | 0.12 | 0.13 | 0.11 | 0.10 | 0.08 | 0.07 | 0.07 | 0.08 | 0.06 | 0.05 | 0.04 |
| **1.0%** | | | | | 1.00 | 0.41 | 0.30 | 0.32 | 0.21 | 0.21 | 0.15 | 0.17 | 0.13 | 0.16 | 0.16 | 0.12 | 0.08 | 0.09 | 0.10 |
| **0.5%** | | | | | | | | 1.00 | 0.88 | 0.60 | 0.59 | 0.58 | 0.52 | 0.53 | 0.46 | 0.41 | 0.28 | 0.42 | 0.33 |

**D**

| Coverage → AF ↓ | 97 | 201 | 237 | 482 | 728 | 969 | 1454 | 1937 | 2413 | 2904 | 3383 | 3872 | 4358 | 4851 | 5855 | 6834 | 7801 | 8790 | 9792 |
|---|---|---|---|---|---|---|---|---|---|---|---|---|---|---|---|---|---|---|---|
| **50.0%** | 3.90% | 4.30% | 2.75% | 1.94% | 1.56% | 1.53% | 1.25% | 1.07% | 0.90% | 0.89% | 0.78% | 0.84% | 0.73% | 0.77% | 0.97% | 0.86% | 0.92% | 0.96% | 0.97% |
| **40.0%** | 5.08% | 5.67% | 3.56% | 2.27% | 1.83% | 1.78% | 1.61% | 1.16% | 0.99% | 0.99% | 0.87% | 0.89% | 0.79% | 0.82% | 1.11% | 1.33% | 1.14% | 0.93% | 0.87% |
| **30.0%** | 5.65% | 6.02% | 3.97% | 2.35% | 1.75% | 1.82% | 1.70% | 1.36% | 1.05% | 1.04% | 0.85% | 0.91% | 0.76% | 0.82% | 1.14% | 0.95% | 0.95% | 1.18% | 1.09% |
| **20.0%** | 4.86% | 6.93% | 4.02% | 2.73% | 1.76% | 1.52% | 1.44% | 1.30% | 1.17% | 1.08% | 0.84% | 0.78% | 0.70% | 0.72% | 0.75% | 0.70% | 0.80% | 0.77% | 0.77% |
| **10.0%** | 3.93% | 5.28% | 3.25% | 2.44% | 1.80% | 1.65% | 1.02% | 0.83% | 0.83% | 0.81% | 0.77% | 0.77% | 0.74% | 0.65% | 0.54% | 0.49% | 0.43% | 0.42% | 0.41% |
| **9.5%** | 3.78% | 5.68% | 3.09% | 2.44% | 1.82% | 1.67% | 0.97% | 0.79% | 0.78% | 0.78% | 0.67% | 0.74% | 0.72% | 0.66% | 0.50% | 0.50% | 0.43% | 0.40% | 0.41% |
| **9.0%** | 3.78% | 5.65% | 2.81% | 2.18% | 1.74% | 1.63% | 0.98% | 0.77% | 0.72% | 0.82% | 0.68% | 0.68% | 0.73% | 0.66% | 0.49% | 0.47% | 0.43% | 0.42% | 0.36% |
| **8.5%** | 3.50% | 5.39% | 2.55% | 2.41% | 1.68% | 1.64% | 0.96% | 0.78% | 0.67% | 0.78% | 0.66% | 0.65% | 0.59% | 0.64% | 0.49% | 0.48% | 0.43% | 0.44% | 0.38% |
| **8.0%** | 3.31% | 5.34% | 2.53% | 2.23% | 1.68% | 1.67% | 1.00% | 0.80% | 0.62% | 0.68% | 0.63% | 0.61% | 0.58% | 0.62% | 0.37% | 0.44% | 0.44% | 0.40% | 0.36% |
| **7.5%** | 3.14% | 4.05% | 2.33% | 1.98% | 1.67% | 1.57% | 1.02% | 0.80% | 0.62% | 0.71% | 0.67% | 0.62% | 0.55% | 0.54% | 0.46% | 0.43% | 0.41% | 0.41% | 0.36% |
| **7.0%** | 3.05% | 4.32% | 2.36% | 2.44% | 1.80% | 1.41% | 1.11% | 0.75% | 0.62% | 0.61% | 0.57% | 0.55% | 0.52% | 0.56% | 0.49% | 0.36% | 0.38% | 0.38% | 0.34% |
| **6.5%** | 2.93% | 4.17% | 2.26% | 2.73% | 1.70% | 1.42% | 1.08% | 0.75% | 0.66% | 0.57% | 0.53% | 0.58% | 0.53% | 0.50% | 0.55% | 0.36% | 0.38% | 0.38% | 0.38% |
| **6.0%** | 2.81% | 4.30% | 2.14% | 2.50% | 1.61% | 1.32% | 1.10% | 0.77% | 0.62% | 0.53% | 0.46% | 0.51% | 0.54% | 0.49% | 0.53% | 0.42% | 0.27% | 0.35% | 0.34% |
| **5.5%** | 2.49% | 2.94% | 2.15% | 2.24% | 1.51% | 1.62% | 1.13% | 0.81% | 0.60% | 0.57% | 0.44% | 0.51% | 0.48% | 0.44% | 0.49% | 0.46% | 0.36% | 0.32% | 0.32% |
| **5.0%** | 2.27% | 1.95% | 2.28% | 2.02% | 1.25% | 1.44% | 1.08% | 0.89% | 0.58% | 0.54% | 0.45% | 0.43% | 0.42% | 0.45% | 0.44% | 0.46% | 0.38% | 0.34% | 0.29% |
| **4.5%** | 2.15% | 1.70% | 2.31% | 1.75% | 1.52% | 1.22% | 0.99% | 0.87% | 0.66% | 0.52% | 0.45% | 0.40% | 0.36% | 0.43% | 0.37% | 0.40% | 0.40% | 0.34% | 0.31% |
| **4.0%** | 1.85% | 1.55% | 1.95% | 1.51% | 1.28% | 1.25% | 0.89% | 0.88% | 0.69% | 0.53% | 0.42% | 0.43% | 0.33% | 0.35% | 0.36% | 0.33% | 0.34% | 0.36% | 0.32% |
| **3.5%** | 1.34% | 1.68% | 1.72% | 1.33% | 1.13% | 1.33% | 1.09% | 0.75% | 0.64% | 0.62% | 0.43% | 0.39% | 0.36% | 0.34% | 0.38% | 0.31% | 0.29% | 0.33% | 0.34% |
| **3.0%** | 1.48% | 1.18% | 1.51% | 1.15% | 0.99% | 1.26% | 0.80% | 0.67% | 0.61% | 0.58% | 0.47% | 0.41% | 0.34% | 0.34% | 0.32% | 0.32% | 0.27% | 0.27% | 0.29% |
| **2.5%** | | | 0.87% | 1.09% | 0.82% | 1.04% | 0.75% | 0.72% | 0.55% | 0.54% | 0.45% | 0.44% | 0.37% | 0.31% | 0.28% | 0.24% | 0.26% | 0.28% | 0.23% |
| **2.0%** | | | 0.96% | 0.99% | 0.71% | 0.74% | 0.81% | 0.65% | 0.54% | 0.45% | 0.43% | 0.44% | 0.35% | 0.36% | 0.28% | 0.22% | 0.24% | 0.23% | 0.22% |
| **1.5%** | | | 0.50% | 0.94% | 0.71% | 0.57% | 0.59% | 0.56% | 0.38% | 0.39% | 0.40% | 0.35% | 0.32% | 0.33% | 0.29% | 0.30% | 0.23% | 0.18% | 0.19% |
| **1.0%** | | | | | 0.55% | 0.48% | 0.37% | 0.36% | 0.34% | 0.37% | 0.31% | 0.30% | 0.25% | 0.28% | 0.18% | 0.21% | 0.24% | 0.19% | 0.22% |
| **0.5%** | | | | | | | | 0.20% | 0.18% | 0.22% | 0.16% | 0.18% | 0.16% | 0.18% | 0.23% | 0.23% | 0.20% | 0.10% | 0.14% |
